# Association of in utero exposure to phthalate and DINCH metabolites with placental DNA methylation

**DOI:** 10.1101/2025.06.26.25330351

**Authors:** Hana Vespalcova, Bethany Knox, Amrit Kaur Sakhi, Cathrine Thomsen, Sofía Aguilar-Lacasaña, Marta Cosin-Tomas, Laura Gómez-Herrera, Olga Sánchez García, Elisa Llurba, María Dolores Gómez-Roig, Jordi Sunyer, Payam Dadvand, Mariona Bustamante, Martine Vrijheid

## Abstract

Phthalates and DINCH are non-persistent chemicals widely used in consumer products. In utero exposure to these compounds has been linked to adverse reproductive and long-term health outcomes, potentially through epigenetic changes in the placenta. This study examined associations between maternal phthalate and DINCH metabolite levels and placental DNA methylation in 469 mother-child pairs from the Barcelona Life Study Cohort (BiSC). Fifteen phthalate and two DINCH metabolites were measured using liquid chromatography–tandem mass spectrometry (LC-MS/MS) in pooled maternal urine samples collected at 19 and 35 weeks of gestation. Placental DNA methylation was assessed using the Illumina EPIC array. We applied robust linear regression for individual metabolite effects and quantile g-computation for mixture analyses. We identified 32 significant associations—24 at 19 weeks and 8 at 35 weeks—across genes enriched in immune and vascular pathways, steroid biosynthesis, and growth factor and sex hormone signaling. Sex-stratified analyses revealed 39 female-specific and 35 male-specific associations, many of which were time-specific. Mixture results aligned with individual metabolite findings. These results suggest that prenatal exposure to phthalates and DINCH may contribute to time- and sex-specific placental epigenetic changes, supporting a role for endocrine disruption and immune modulation in fetal development.

**Highlights:**

- In utero exposure to phthalates and DINCH was associated with differential DNA methylation in the placenta
- Associations were dependent of sex and in utero exposure window
- Identified genes were involved in immune and vascular regulation, steroid biosynthesis, and signaling through growth factors and sex hormone receptors

## 1. Introduction

Phthalates and 1,2-cyclohexane dicarboxylic acid diisononyl ester (DINCH), non-persistent chemicals, are commonly used plasticizers and stabilizers typically added to plastic materials such as food packaging, cosmetics, toys or drug films and other medical devices^1–3^. Due to their widespread use, phthalates and DINCH have become ubiquitous in the environment, leading to chronic exposure through ingestion, inhalation, and skin absorption in humans^1,4^.

Exposure to phthalates has been linked to a broad range of health disorders^5,6^. In particular, the in utero exposure has been related to an increased risk of pregnancy complications such as miscarriage, preeclampsia, or gestational diabetes^7–9^, and also to short and long-term health consequences in the offspring, including intrauterine growth restriction, preterm birth, low birth weight, obesity, immune disorders or neurodevelopmental disorders ^10,11^. Some of these associations have been shown to be sex-specific, such as reduced ovarian weight or altered puberty timing in females, and altered testosterone synthesis or increased risk of ADHD in males, among others^12–15^.

During pregnancy, the placenta is an essential organ for fetal growth and development. It supports nutrient and oxygen exchange, immune defense and endocrine regulation, while fetal organs are developed^16^. The placenta is permeable to phthalates and DINCH, with several studies reporting comparable levels of these substances in both maternal and cord blood^17–21^. Phthalates and DINCH can deregulate different biological processes^22,23^, but they are particularly known for their endocrine disrupting effects^24^. Therefore, phthalates and DINCH may impair placental formation and function, either directly or indirectly by endocrine disruption in pregnant women. These alterations in placental formation and function can be recorded in the placental epigenome^25^. The epigenome refers to the collection of chemical changes to DNA and its associated proteins, including DNA methylation, histone modifications, and changes in non-coding RNA expression, all of which influence gene expression without altering the underlying DNA sequence^26,27^.

Several studies have investigated the association of in utero phthalate and DINCH exposure with epigenetic alterations in the offspring. In cord blood, epigenome-wide association studies (EWAS), ranging from 64 to 336 samples, have reported up to 25 CpGs with differential DNA methylation^28–34^. In the placenta, two EWAS studies with 202 and 387 samples from two French cohorts have identified 1 and 114 differently methylated CpGs, respectively^35,36^. However, these studies were relatively small, and some of them did not explore the whole genome, or the effects of the exposure window or fetal sex. Moreover, assessment of in utero phthalate and DINCH exposure was based on one or a few spot urine samples in all but one study^36^, potentially producing high levels of measurement error, since exposure to these short-lived chemicals is known to be highly variable.

In this study we aimed to evaluate the association between phthalate and DINCH metabolite levels measured in pooled maternal urine samples collected at 19 and 35 weeks of gestation, and placental genome-wide DNA methylation in 469 mother-child pairs from the Barcelona Life Study Cohort (BiSC).

## 2. Material and methods

### 2.1 Study population

This study was nested in the Barcelona Life Study Cohort (BiSC), a population-based birth cohort in Barcelona, Spain^37^. Pregnant women (n=1,080) were recruited during the first trimester of pregnancy between 2018 and 2021 in three hospitals in Barcelona (Hospital Sant Joan de Déu, Hospital de la Santa Creu i Sant Pau, and Hospital Clínic de Barcelona). The detailed information about inclusion criteria, follow-up visits and data collected can be seen elsewhere^37^. Briefly, we included pregnant women between 18 and 45 years old, living in the catchment area of recruiting hospitals and being able to communicate in Catalan, Spanish, or English. We excluded women having a fetus with major congenital anomalies (i.e., malformations incompatible with life). In this study we examined 469 BiSC mother-child pairs with available data of urinary phthalate and DINCH levels in 19 weeks of gestation (19w, N=349) or 35 weeks of gestation (35w, N=404) of pregnancy (overlap 284), placental DNA methylation, gestational age, and covariates (Figure S1).

All participants signed an informed consent form prior to their enrolment into the cohort. Ethics approvals were obtained from the corresponding authorities in all the participating institutions and hospitals. Study data were collected and managed using REDCap electronic data capture tools hosted at ISGlobal^38,39^.

### 2.2 Exposure assessment

Pregnant participants’ first and last voids of the day were collected during six consecutive days at approximately 19w (mean (standard deviation (SD))=18.8 (2.9) weeks, N=695 samples) and 35w (mean (SD)=34.7 (1), N=756 samples). The samples were kept in freezers of participants’ home during the sampling week and then transported to the BiSC biobank and kept in −80°C freezers till the time of the analysis. Urine pools, containing equivalent volumes of each void (with a minimum of 10 voids), were created for each participant at each collection time point.

A total of 15 phthalate metabolites and 2 DINCH metabolites were measured in each urine pool using high-performance liquid chromatography and tandem mass spectrometry (LC-MS/MS) at the Norwegian Institute of Public Health^40^. These included: monoethyl phthalate (MEP), mono-iso-butyl phthalate (MiBP), mono-n-butyl phthalate (MnBP), mono benzyl phthalate (MBzP), mono-n-pentyl phthalate (MnPeP), monocyclohexyl phthalate (MCHP), mono-n-octyl phthalate (MnOP), mono-2-ethylhexyl phthalate (MEHP), mono-2-ethyl-5-hydroxyhexyl phthalate (MEHHP), mono-2-ethyl-5-oxohexyl phthalate (MEOHP), mono-2-ethyl 5-carboxypentyl phthalate (MECPP), mono-4-methyl-7-hydroxyoctyl phthalate (oh-MiNP), mono-4-methyl-7-oxooctyl phthalate (oxo-MiNP), mono-4-methyl-7-carboxyoctyl phthalate (cx-MiNP), 6-hydroxy monopropylheptylphthalate (oh-MPHP), 2-(((hydroxy-4-methyloctyl)oxy)carbonyl)cyclohexanecarboxylic acid (oh-MINCH), 2-(((4-methyl-7-oxyooctyl)oxy)carbonyl)cyclohexanecarboxylic acid (oxo-MINCH). Some of the phthalate metabolites (oh-MiNP, oxo-MiNP, cx-MiNP and oh-MPHP) and DINCH metabolites (oh-MINCH and oxo-MINCH) have multiple isomers. Thus, for these metabolites the sum of the isomers was reported. Three blanks and two in-house control samples were analyzed alongside the urine samples in each laboratory batch (about 50 samples per batch). For MBzP, MEHP, MEHHP and MEOHP, a signal was detected in <25% of the analyzed blanks with a minimal signal (<2 times limits of quantification), thus no correction was indicated. For the remaining compounds no blank detection was observed. The coefficient of variation of control samples within batches ranged between 7-33%. Phthalate and DINCH levels are expressed as ng/mL urine.

oh-MiNP was discarded from the analysis due to co-elution with a metabolite of a terephthalate that has the same mass and retention time (mono-2-ethyl-5-carboxypentyl terephthalate, MECPTP). Three phthalate metabolites, MnPeP, MCHP, MnOP were detected in <5% of samples and were also excluded from the analysis. The limit of detection (LOD), the lower and upper limit of quantification (LLOQ, ULOQ), and the descriptive of the subset of samples examined in this study (19w: N=349, 35w: N=404) are shown in Tables TS1A-B. After testing for normal distribution, phthalate levels were log2-transformed to meet the requirement of Gaussian distribution for analysis. For phthalates where no levels were detected due to signals <LOD, the missing values were imputed using the left-censored ‘fill_in’ imputation method described by Helsel (1990)^41^. Additionally, the weighted sum of metabolites (nmol/L) derived from the same parent compound was calculated by summing the metabolite levels (in original scale and with values <LOD imputed) divided by their molecular weight (sum DEHP: MEHP, MEHHP, MEOHP and MECPP, sum DiNP: oxo-MiNP and cx-MiNP, sum DINCH: oh-MINCH and oxo-MINCH). Then, the sums were tested for normal distribution and log2-transformed.

The effect of several technical and design variables (laboratory batch, urine time to storage, number of urine samples in the pool), and biological variables (urine creatinine levels and gestational age) on phthalate levels were investigated, and found to be minimal with the exception of creatinine. Creatinine levels (ml/l urine) were measured in the urine pool as described elsewhere^42^. To eliminate this potential noise, we applied a standardization method, based on linear regression that adjust the exposure for creatinine protecting other variables (hospital of delivery, COVID19 confinement, gestational age, maternal smoking, education, BMI and age, child’s ethnicity and sex, and parity) described by Mortamais, et al. (2012)^43^. This second dataset corrected for creatinine has been used in the sensitivity analysis (see section 2.5).

### 2.3 DNA methylation

#### 2.3.1 Placental biopsy collection

Following a harmonized protocol across hospitals, placental biopsies (2.5 × 1 cm, N = 611) were collected from two opposite quadrants approximately 3–4 cm from the cord insertion site. Biopsies were then halved (2.5 × 0.5 cm). One half was stored in liquid nitrogen, and the other half treated with RNAlater and dissected into four 0.5 × 0.5 cm samples representing the chorionic plate/membranes, upper chorionic villi, lower chorionic villi, and decidua. All biopsies were finally stored at −80°C until the time of analysis.

#### 2.3.2 Placental DNA extraction

The DNA was extracted from 30-40 mg of the upper chorionic villi collected in RNAlater. First, the dissected samples were homogenized using a bead mill (bead beater) at 4°C for 26 seconds. Next, the genomic DNA was isolated using the AllPrep®DNA/RNA/miRNA Universal Kit, (Qiagen, CA, USA). DNA was eluted in 80 and 60 µl and stored in different aliquots at −80°C. DNA quality was evaluated on a NanoDrop spectrophotometer (Thermo Scientific, Waltham, MA, USA) and additionally 500 ng of DNA was run on 1% agarose gels to confirm that samples did not present visual signs of degradation.

#### 2.3.3 Placental DNA methylation data acquisition

DNA methylation analysis was performed on 624 DNA samples from 589 individuals including 35 duplicates. Seven hundred and fifty ng of DNA were bisulfite-converted using the EZ 96-DNA methylation kit following the manufacturer’s standard protocol. The DNA methylation analysis was conducted using the Infinium Methylation EPIC BeadChip from Illumina, following manufacturer’s protocol in the Human Genome facility (HUGE-F) at the Erasmus Medical Centre core facility. The samples were randomly distributed into multiple plates, and the random distribution according to main biological and socioeconomic factors was checked.

#### 2.3.4 Placental DNA methylation data quality control and normalization

The DNA methylation data pre-processing was performed using the PACEAnalysis R package (v.0.1.9) (https://www.epicenteredresearch.com/). During the data pre-processing, sample quality control, probe quality control, normalization, batch correction, estimation of cell type proportions, and winsorization of outlier values were conducted. Overall, 59 samples were excluded from the data due to: a) low quality (N=5), b) sample call rate < 95% (N=3), c) sex inconsistencies (N=9), d) substantial contamination with DNA from other samples (log2 odds < −1) (N=11)^44^, e) duplicates calculated using the probes to genotype 59 single nucleotide polymorphisms (SNPs) contained in the array (N=29), and f) siblings (N=2). The 59 excluded samples correspond to 24 individuals. After sample quality control, 565 unique samples remained. Probes with a call rate < 95% were discarded according to the SeSAMe method^45^. Moreover, control and non-CpG probes, sex chromosome targeting probes, probes giving inconsistent results between 450K and EPIC arrays^46^, and problematic probes (probes with hybridizing problems and probes affected by the presence of SNPs: probes containing a SNP at the CpG site itself, probes with a SNP adjacent to the CpG site, probes with a SNP within 5 nucleotides of the CpG site if the minor allele frequency (MAF) was greater than 1% in global populations) according to Zhou et al. (2017)^45^, were excluded.

The dye-bias and Noob background correction, and normalization with the functional normalization method were conducted using the minfi R package^47,48^. Correction for type 2 probe bias was performed applying the beta-mixture quantile normalization (BMIQ)^49^. Data was explored using Principal Component Analysis (PCA) and associations of the first 12 PCs with main variables (hospital of recruitment, hospital of delivery, sex, ethnic, covid confinement, covid pandemic, gestational age, birth weight, maternal smoking) and technical variables (array, working plate, DNA extraction batch, round of extraction, DNA concentration, quality ratio 260/280 and quality ratio 260/230) were examined using linear regression models. The effect of the array batch variable was eliminated using the ComBat method due to its association with most of the PCs, including PC1, which explained 30.36% of the variance^50^. DNA methylation values are expressed as beta values, where 0 indicates no methylation and 1 indicates complete methylation. The beta values were winsorized based on a percentile of 1% (0.5% on each side) estimated from the empirical beta-distribution to reduce the influence of outliers.

### 2.4 Covariates

Based on evidence from previous studies, the following variables were included as covariates in the models. Maternal age (years), maternal education (primary and secondary school/university), maternal tobacco smoking status during pregnancy (non-smoker/smoker), parity (multiparous/nulliparous), and child’s ethnicity (European/other) were obtained from self-reported questionnaires. Maternal body mass index (BMI) at 12 weeks of pregnancy (kg/m^2^) was obtained through direct measurements. Gestational age at delivery (weeks) was calculated based on the crown-rump-length, measured using ultrasound examination at approximately 12th gestational week obstetric visit (22). Placental cell type proportions were estimated from the DNA methylation data (see above). Child’s sex (female/male), and birth weight (grams), the later used in the descriptive of the study, were obtained from medical records. COVID-19 confinement (whether pregnancy took place before confinement/during confinement/after confinement) and hospital of delivery (Hospital Sant Joan de Déu, Hospital de la Santa Creu i Sant Pau, and other) were obtained based on conception and gestational age and the hospital where the delivery took place.

### 2.5 Statistical analyses

Prior to the main analysis, we calculated the Spearman’s correlation between phthalate and DINCH metabolites and the Spearman’s correlation with creatinine in each trimester. To compare the variability of phthalate levels between trimesters, we calculated the intra-class (ICC) that calculates within-group variation (ie. within the same mother).

The association of in utero exposure to phthalates and DINCH at different exposure windows (19w and 35w) with placental DNA methylation was analyzed using robust linear regression models for each exposure and CpG site (*PACEanalysis* R package). The main model was adjusted for hospital of delivery, COVID19 confinement, maternal smoking, maternal education, maternal BMI at 12 weeks of pregnancy, maternal age at delivery, parity, ethnicity and sex of the offspring, and placental cell composition.

To control for multiple-testing the Bonferroni correction was applied considering the total number of CpGs tested for each metabolite and time point (a p value <1E-07 was considered statistically significant). The lambda inflation factor was calculated as a ratio between median of observed distribution of p-values and median of expected distribution of p-values^51^. Effect size was expressed as the change in DNA methylation (from 0 to 1) per doubling in phthalate levels (ng/mL).

To better reflect the actual exposure in the presence of other compounds, we repeated EWAS on the sum of metabolites (sum DiNP, sum DEHP and sum DINCH). Additionally, we performed the analysis of chemical mixtures using quantile g-computation implemented in *qgcomp* R package to examine the potential mixture effect of phthalate metabolites on DNA methylation^52^. The quantile g-computation estimates the overall and partial effects of mixture exposure on the outcome of interest using quantiles of metabolite levels, while allowing for covariate adjustment. This analysis was restricted to statistically significant CpGs observed from the EWAS of single metabolites.

We further stratified the analyses by sex. Differences in the effect size of Bonferroni significant CpGs between sexes were compared with the Cochran’s *Q* test implemented in *meta* R package^53^. The p-value of the Q test was adjusted by the number of associations identified in boys and girls (p-value threshold <7E-04).

Finally, a number of sensitivity analyses were conducted: a) additionally adjusting for gestational age, which may be a mediator in the associations; b) restricting the analyses to the subset of children of European origin, a more homogeneous population at the genetic level; c) not adjusting for placental cellular composition, to test whether epigenetic associations were driven by true methylation changes rather than shifts in cell proportions; and d) using the phthalate levels corrected for creatinine, in order to account for variability in urine dilution in pools.

### 2.6 Downstream analyses

The IlluminaHumanMethylationEPICanno.ilm10b4.hg19 R package^54^ was used to annotate the CpGs to genes (within the gene or at <1,500 bp upstream of the gene start site) and to CpG islands. In addition, to annotate the CpGs to enhancers and link them to genes in placenta tissue, we used the EPIraction webpage tool (run 14/08/2024)^55^. Furthermore, we searched whether identified CpGs overlapped with placental methylation quantitative trait loci (mQTLs), placental germline differentially methylated regions (gDMRs)^56^, placental partially methylated domains (PMDs) and placental chromatin states according to ROADMAP^57^. Placental fetal cis-mQTLs were identified in BiSC (for more information see Supplementary Methods).

To identify potential biological pathways underlying the effects of identified DNA methylation changes (suggestive CpGs at p value <1E-05), we conducted several functional enrichment analyzes using the missMethyl R package^58^. We tested enrichment for: (i) gene ontologies (GO); (ii) biological pathways (KEGG, Wiki Pathways, Reactome, BioCarta, Panther, MsigDB); (iii) transcription factors (ENCODE, ChEA); and (iv) diseases (DisGeNET, DsigDB, OMIM Diseases, OMIM Expanded, GWAS Catalogue). A minimum of two genes in the pathway and a False Discovery Rate (FDR) <0.2 was needed to report the pathways in the manuscript. Finally, we searched for previous associations with traits and exposures of significant CpGs in the EWAS Atlas^59^, EWAS catalog^60^, and the Comparative Toxicogenomics Database (CTD)^61^.

All analyses were performed using R Statistical Software (v4.1.2, v4.2.2, and v4.4.3)^62–64^.

## 3. Results

### 3.1 Study population

Our study sample included 469 pregnant women and their offspring from the BiSC cohort, 349 with phthalate levels measured at 19w (mean: 18.74, SD: 2.86), and 404 at 35w (mean: 34.74, SD: 1.38), with an overlap of 284. The average maternal age at the enrollment was 34.6 years and the average BMI at 12w was 24.5 kg/m^2^. Overall, most of the women had completed university education (69%) and did not smoke during pregnancy (92%). Around 23% of participants were pregnant during the COVID-19 confinement and half (50%) delivered in Hospital Sant Joan de Déu and 46% in Hospital Sant Pau. The participants gave birth at an average gestational age of 39.8 weeks and 55% of them were nulliparous. Average birth weight was 3287.4 g, the sex of the babies was uniformly distributed and the majority of the babies were of European origin (70%). Detailed information of the participants is provided in Table 1.

**Table 1.** Descriptives of the study population. N: sample size; SD: standard deviation

| **Table 1. Descriptives of the study population** |  |  |  |  |  |  |
| --- | --- | --- | --- | --- | --- | --- |
| N: sample size; SD: standard deviation |  |  |  |  |  |  |
|  | **Whole population**<br>**(N=469)** |  | **Population at 19w**<br>**(N=349)** |  | **Population at 35w**<br>**(N=404)** |  |
|  | **N** | **%** | **N** | **%** | **N** | **%** |
| **Hospital of delivery** |  |  |  |  |  |  |
| Hospital Sant Pau | 217 | 46 | 158 | 45 | 195 | 48 |
| Hospital Sant Joan de Déu | 235 | 50 | 182 | 52 | 194 | 48 |
| Other | 17 | 4 | 9 | 3 | 15 | 4 |
| **Covid-19 confinement** |  |  |  |  |  |  |
| Pregnancy before confinement | 210 | 45 | 130 | 37 | 190 | 47 |
| Pregnancy during confinement | 109 | 23 | 89 | 26 | 82 | 20 |
| Pregnancy after confinement | 150 | 32 | 130 | 37 | 132 | 33 |
| **Maternal education** |  |  |  |  |  |  |
| Primary or secondary school | 143 | 30 | 105 | 30 | 119 | 29 |
| University | 326 | 70 | 244 | 70 | 285 | 71 |
| **Maternal smoking during pregnancy** |  |  |  |  |  |  |
| Non-smoker | 434 | 93 | 323 | 93 | 376 | 93 |
| Smoker | 35 | 7 | 26 | 7 | 28 | 7 |
| **Parity** |  |  |  |  |  |  |
| Multiparous | 207 | 44 | 161 | 46 | 179 | 44 |
| Nulliparous | 262 | 56 | 188 | 54 | 225 | 56 |
| **Child ethnicity** |  |  |  |  |  |  |
| European | 329 | 70 | 243 | 70 | 292 | 72 |
| Other | 140 | 30 | 106 | 30 | 112 | 28 |
| **Child sex** |  |  |  |  |  |  |
| Female | 238 | 51 | 181 | 52 | 212 | 52 |
| Male | 231 | 49 | 168 | 48 | 192 | 48 |
|  | **Mean** | **SD** | **Mean** | **SD** | **Mean** | **SD** |
| **Maternal age (years)** | 34.55 | 4.55 | 34.63 | 4.5 | 34.7 | 4.37 |
| **Maternal body mass index (BMI) at 12 weeks of pregnancy (kg/m<sup>2</sup>)** | 24.53 | 4.44 | 24.51 | 4.39 | 24.33 | 4.31 |
| **Gestational age at birth (weeks)** | 39.75 | 1.4 | 39.68 | 1.42 | 39.85 | 1.22 |
| **Birth weight (g)** | 3287.42 | 465.65 | 3281.88 | 492.92 | 3309.06 | 429.68 |
| **Cell type proportions** |  |  |  |  |  |  |
| Trophoblasts | 0.08 | 0.07 | 0.07 | 0.06 | 0.07 | 0.07 |
| Stromal | 0.01 | 0.01 | 0.01 | 0.01 | 0.01 | 0.01 |
| Hofbauer | <0.01 | 0.01 | <0.01 | 0.01 | <0.01 | 0.01 |
| Endothelial | 0.05 | 0.03 | 0.05 | 0.03 | 0.05 | 0.03 |
| Nucleated red blood cells (nRBC) | 0.02 | 0.02 | 0.01 | 0.02 | 0.02 | 0.02 |
| Syncytiotrophoblast | 0.84 | 0.1 | 0.84 | 0.1 | 0.84 | 0.1 |

### 3.2 Exposure to phthalates and DINCH

Eleven phthalate and 2 DINCH metabolites with levels >LOD in more than 90% of the samples were included in the analyses. Median and interquartile range (IQR) levels for each compound and period are presented in Table 2 (see TS2A, TS2B for more details). MEP had the highest levels measured in both periods of pregnancy (median 19w: 47.51 ng/mL, 35w: 40.04 ng/mL). The weighted sums of metabolites of DEHP (4 metabolites), DiNP (2 metabolites), and DINCH (2 metabolites) showed median levels of 67.71, 40.51 and 10.19 nmol/L at 19w, and 45.23, 25.56, 6.13 nmol/L at 35w, respectively (Table 2, TS2A, TS2B).

**Table 2.** Descriptives of the phthalate and DINCH metabolite levels in maternal urine samples collected at 19w (N=349) and 35w (N=404) Only the 13 phthalate and DINCH metabolites (and the sums) included in this study are reported. N: sample size Units: ng/mL urine IQR: interquartile range

| Variable | Name | 19w | 35w |
| --- | --- | --- | --- |
|  |  | Median (IQR) | Median (IQR) |
| MEP | Monoethyl phthalate | 47.51 (22.16-89.09) | 40.04 (21.03-92.27) |
| MiBP | Mono-iso-butyl phthalate | 11.58 (7.50-17.95) | 11.38 (7.14-18.15) |
| MnBP | Mono-n-butyl phthalate | 10.51 (6.73-15.50) | 9.81 (6.59-16.91) |
| MBzP | Mono benzyl phthalate | 1.24 (0.75-2.15) | 1.16 (0.59-2.13) |
| MEHP | Mono-2-ethylhexyl phthalate | 1.82 (1.05-3.11) | 1.38 (0.68-2.71) |
| MEHHP | Mono-2-ethyl-5-hydroxyhexyl phthalate | 5.68 (3.85-8.30) | 5.06 (3.54-8.68) |
| MEOHP | Mono-2-ethyl-5-oxohexyl phthalate | 4.71 (3.13-7.00) | 4.61 (3.07-7.25) |
| MECPP | Mono-2-ethyl 5-carboxypentyl phthalate | 8.52 (6.31-12.11) | 8.29 (5.79-12.64) |
| oxo-MiNP | Mono-4-methyl-7-oxooctyl phthalate | 4.31 (2.81-7.32) | 4.32 (2.627-3.33) |
| cx-MiNP | Mono-4-methyl-7-carboxyoctyl phthalate | 7.97 (5.51-11.76) | 7.02 (5.08-11.02) |
| oh-MPHP | 6-Hydroxy Monopropylheptylphthalate | 1.81 (1.17-2.99) | 1.87 (1.13-3.29) |
| oh-MINCH | 2-(((Hydroxy-4-methyloctyl)oxy)carbonyl)cyclohexanecarboxylic acid | 1.94 (1.33-3.37) | 1.71 (1.10-3.13) |
| oxo-MINCH | 2-(((4-Methyl-7-oxyooctyl)oxy)carbonyl)cyclohexanecarboxylic acid | 1.21 (0.73-2.08) | 1.19 (0.71-2.19) |
| sum DEHP | molar sum of MEHP, MEHHP, MEOHP and MECPP (nmol/L) | 67.71 (50.00-99.75) | 63.75 (45.23-102.30) |
| sum DiNP | molar sum of oxo-MiNP and cx-MiNP (nmol/L) | 40.51 (28.22-63.59) | 37.59 (25.55-60.12) |
| sum DINCH | molar sum of oh-MINCH and oxo-MINCH (nmol/L) | 10.19 (6.61-17.78) | 9.66 (6.13-17.81) |

The metabolites from different parent compounds presented a moderate to weak correlation, with the highest correlation between oh-MPHP and the oxo-MiNP (Spearman’s correlation coefficient of 0.59 at 19w, and 0.61 at 35w) (Figures 1A, 1B, Figure S2A, S2B). The correlation within metabolites from the same parent compound was higher, ranging from 0.73 to 0.95 at 19w and 0.70 to 0.95 at 35w (Figures 1A, 1B). The intra-class correlation of 19w and 35w ranged between 0.29 and 0.58 (Figure S3), indicating moderate correlation within the same women between trimesters. Finally, the metabolite levels at 19w and 35w were moderately correlated with creatinine (Spearman’s correlation coefficient at 19w: 0.28-0.43, 35w: 0.30-0.46) (Figures S4A, S4B).

**Figure 1.**
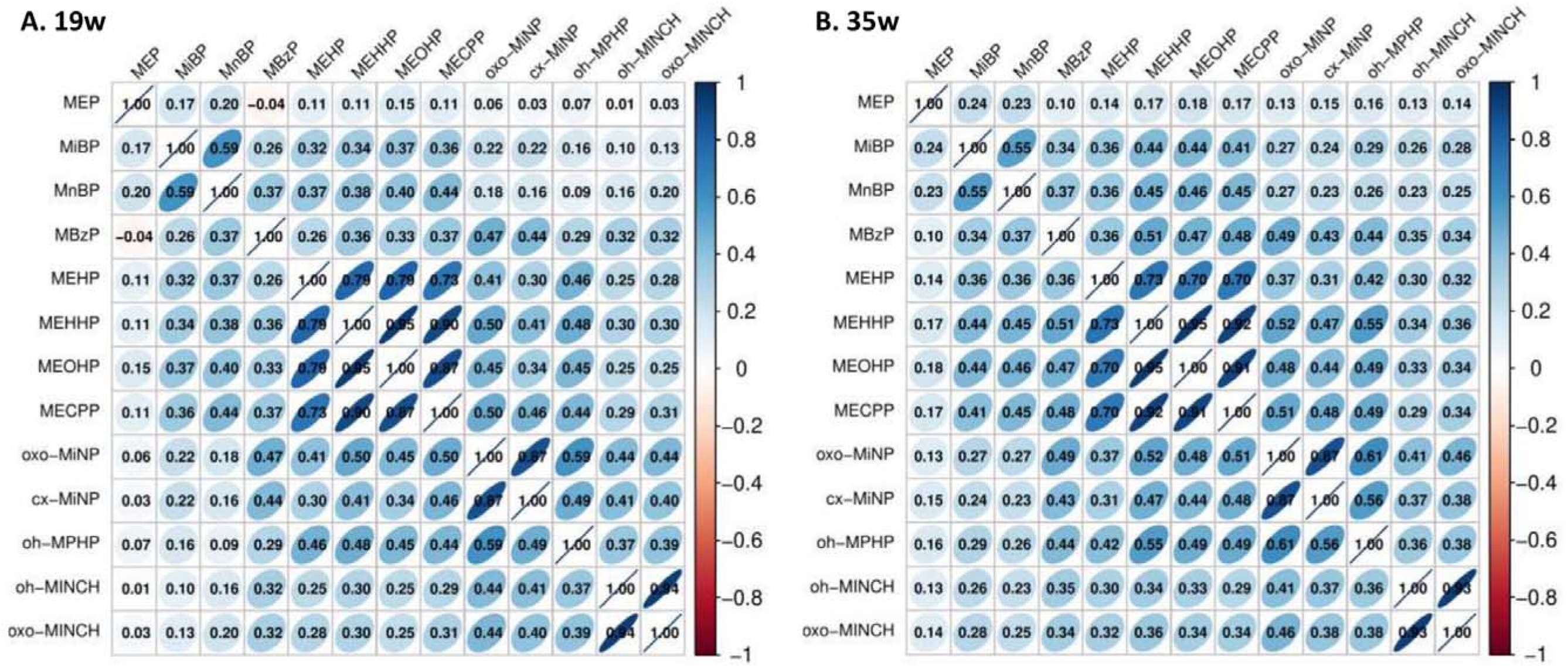
Correlation of phthalate and DINCH metabolite levels. The heatmap shows the Pearson’s correlation of the 13 phthalate and DINCH metabolite levels measured at 19w (A) and at 35w (B). Positive correlations are indicated in blue and inverse correlations in red.

### 3.3 Epigenome-wide association study by exposure period

In the main analysis of phthalates and DINCH metabolites and placental DNA methylation, a total of 32 associations were identified after Bonferroni correction (p value <1E-07), and 788 at suggestive statistical significance (p value <1E-05) (Figure 2, Table S3). The genomic inflation factors (λ) ranged from 0.95 to 1.22 at 19w, and from 0.84 to 1.27 at 35w (Table S3).

**Figure 2.**
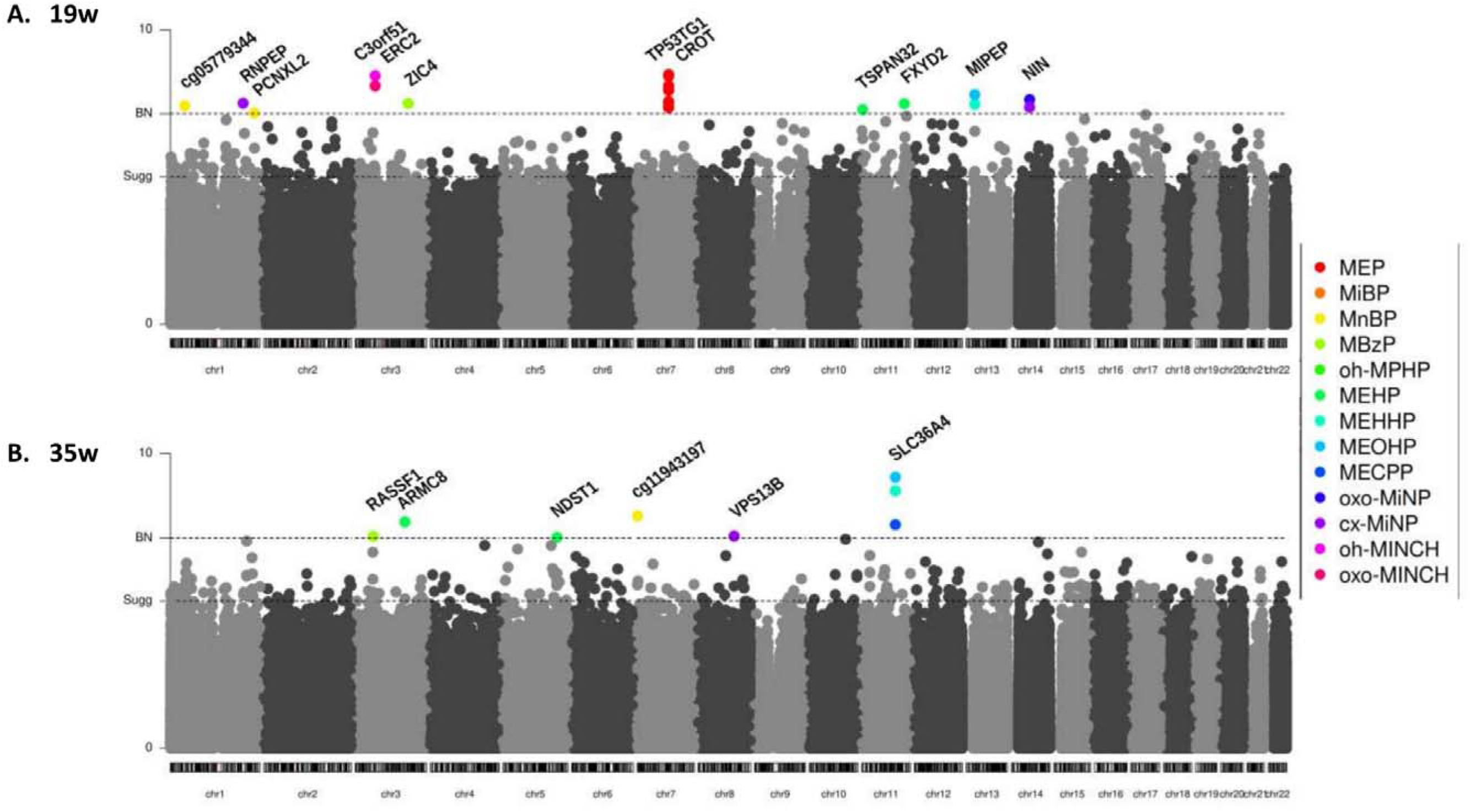
Manhathan plots of the association between genome-wide DNA methylation and phthalate and DINCH metabolite levels, by exposure period. Each dot is the association of the DNA methylation levels at one CpG with the levels of one of the phthalate or DINCH metabolites measured at 19w (A) and at 35w (B). The x-axis represents the position of the CpGs and the y-axis the -log10 p value of the association. The horizontal dashed lines are the suggestive (1E-05 p value <1E-05) and Bonferroni (p value <1E-7) multiple-testing thresholds. Associations surviving Bonferroni correction are coloured according to the legend.

In particular, at 19w, 24 Bonferroni-adjusted significant associations were found, which represented 10 unique metabolites and 21 unique CpGs, distributed in 10 loci (defined as a region <1 Mb) and annotated to 11 unique genes (Table S4). Twelve CpGs from the same locus at chromosome 7 (cg12969170 as the lead CpG) were inversely associated with MEP levels. The rest of CpGs represented different loci and were associated with one or a few metabolites of the same parental compound (Figure S5A). The effect sizes of the significant CpGs ranged from −0.047 to 0.013 for a doubling in the exposure levels, with most of them (82%) showing decreased methylation levels with increasing exposure levels (Table S4). In general, DNA methylation levels at significant CpGs presented the same direction of effects across compounds with some exceptions. For instance, the inverse associations between MEP and CpGs at chromosome 7 were in the same direction in most of the compounds, but MiBP, MnBP or MBzP, which showed increased levels of methylation at these CpGs (Figure S5A). Suggestive associations (N=493) are shown in Table S5A.

At 35w, we identified 8 Bonferroni-adjusted significant associations, representing 7 unique metabolites and 6 unique CpGs (6 loci, 5 unique genes) (Table S4). The effect size of significant CpGs ranged from −0.011 to 0.017, with 63% of the associations being positive (Figure S5B). Similar to the 19w exposure period, the CpGs showed similar effects across compounds and there was some overlap among the metabolites of the same parental compound (Figure S5B). Suggestive associations (N=295) are shown in Table S5B.

There was no overlap of Bonferroni significant metabolite-CpG pairs between each exposure period. Moreover, some of significant associations in one exposure period showed week effect sizes in the other, suggesting period-specific effects (Figure 3). For instance, statistically significant CpGs that were associated with exposure to MEP metabolite at 19w showed a weak effect at 35w, while cg09012471 associated with MEHP metabolite at 35w showed no effect at 19w.

**Figure 3.**
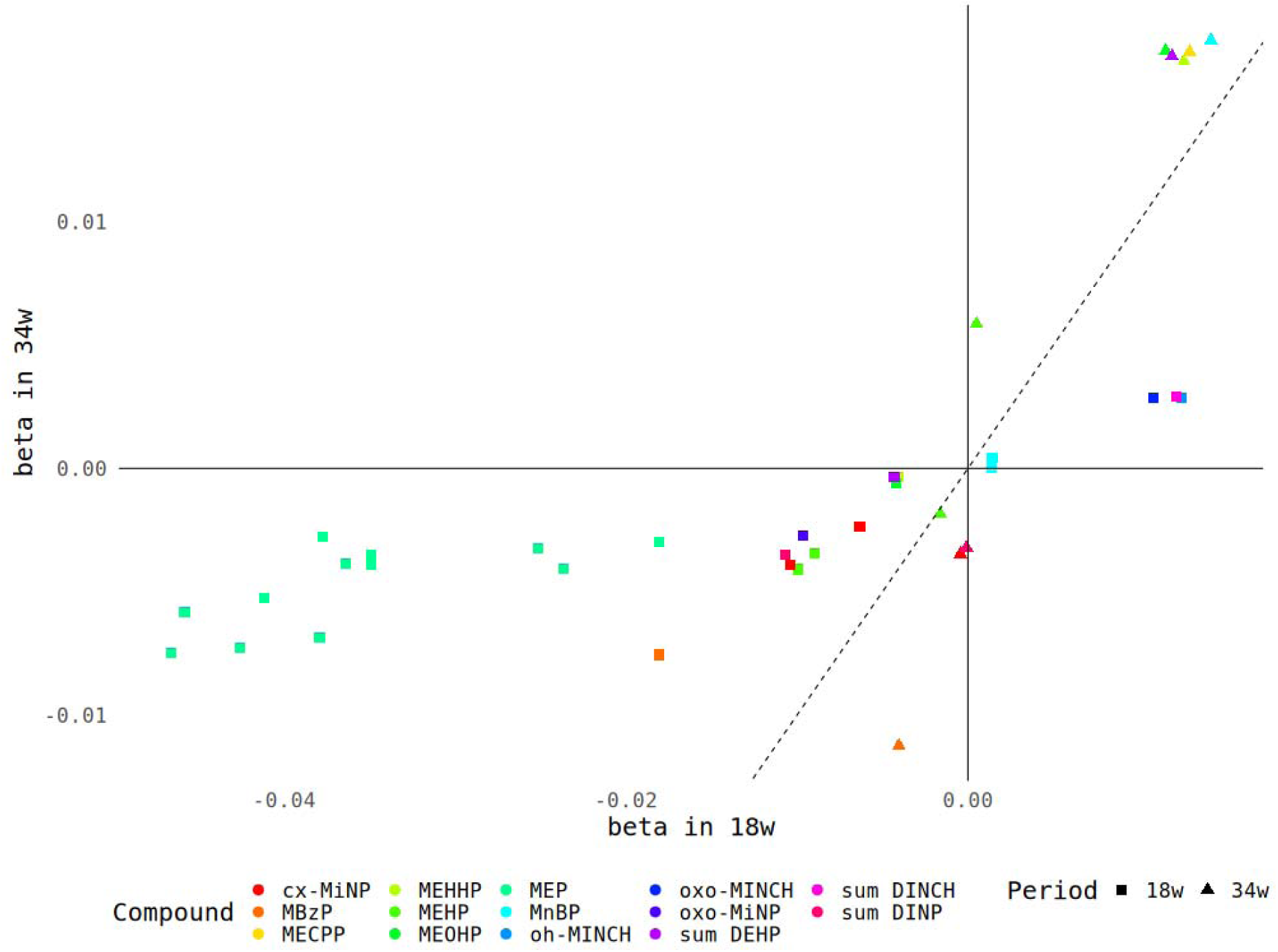
Correlation of the effect sizes of the Bonferroni significant CpGs between exposure periods (19w and 35w) Each symbol is the association of the DNA methylation levels at one CpG with the levels of one of the phthalate or DINCH metabolites. The x-axis represents the effect size of the association with the exposure levels measured at 19w, and the y-axis at 35w.The shape of the symbols represents the period when the association is significant, and the color the phthalate or DINCH metabolite (see legend). Symbols at the diagonal indicate similar effect sizes, while at the axes indicate period-specific effects.

### 3.4 Sum of metabolites and mixture analyses

The results for the sum of metabolites were consistent with the associations observed for individual compounds at both exposure windows. (Tables S6, S7, S8). We also conducted mixture analyses for the Bonferroni-significant CpGs, identifying associations for eight of the 27 significant CpGs from the single-metabolite analysis at 19 weeks (Table S9A), and for three of the seven CpGs at 35 weeks, after correcting for multiple testing (Table S9B). The overall effect sizes of the associations between the mixture and these CpGs ranged from −0.430 to 0.025, and the direction of the effects corresponded to the direction of significant associations observed for the individual metabolites. The partial effects of the metabolites on differently methylated CpGs in the mixture analysis are shown in Figures S6.

### 3.5 Epigenome-wide association analysis stratified by sex

Results of the sex-stratified analyses by exposure period are shown in Tables S10A and S10B. After applying the Cochran’s *Q* heterogeneity test, 43 associations showed evidence of differences in effect sizes between females and males at 19w, and 31 associations at 35w. The associations at 19w included 23 unique CpGs (20 loci, 19 unique genes) and 10 unique metabolites in females, and 17 unique CpGs (17 loci, 17 unique genes) and 10 unique metabolites in males (Tables S11, S12, S13). The associations at 35w included 14 CpGs (14 loci, 9 unique genes) and 8 metabolites in females, and 16 unique CpGs (13 loci, 10 unique genes) and 7 unique metabolites in males (Tables S11, S12, S13). The effect sizes of the sex-specific CpGs across sexes are shown in Figure 4.

**Figure 4.**
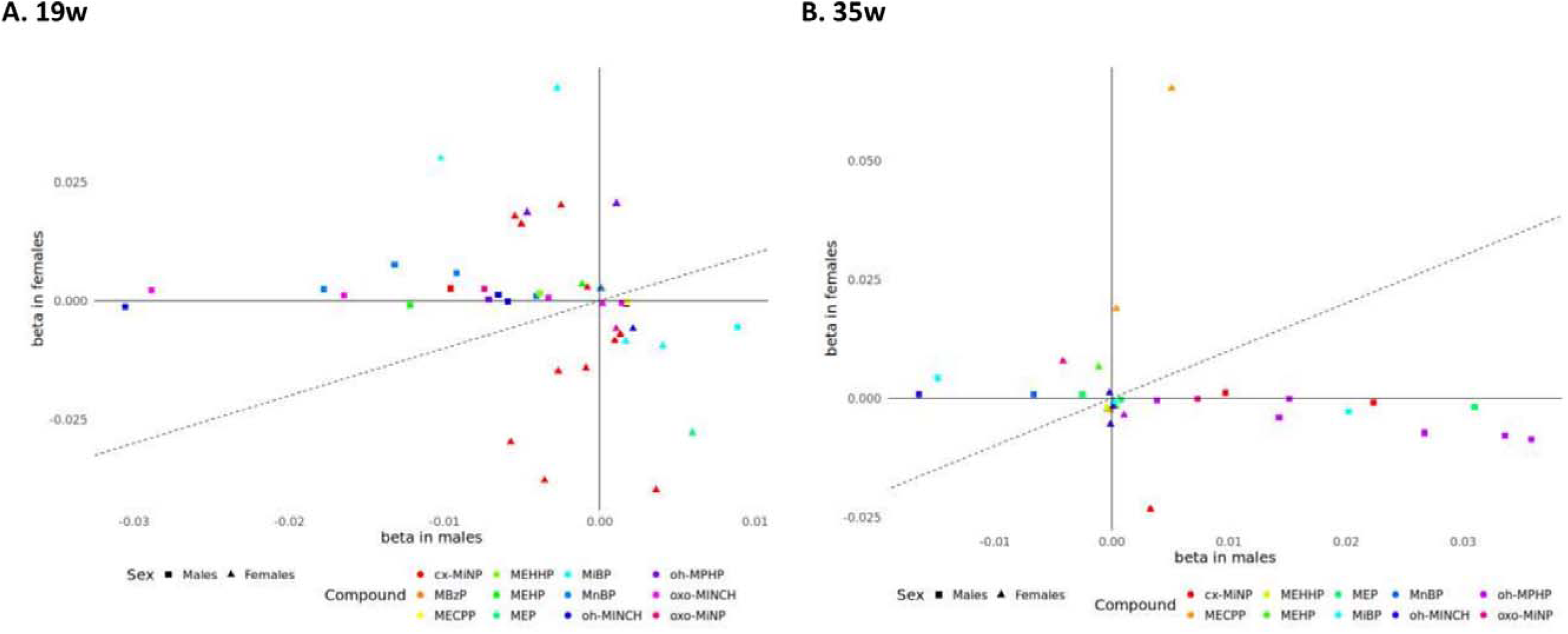
Correlation of the effect sizes of the Bonferroni significant CpGs between males and females, by exposure periods. Each symbol is the association of the DNA methylation levels at one CpG with the levels of one of the phthalate or DINCH metabolites measured at 19w (A) and at 35w (B). The x-axis represents the effect in males, and the y-axis the effect in females. The shape of the symbols represents the sex for which the CpG is significant, and the color the phthalate or DINCH metabolite (see legend). Symbols at the diagonal indicate similar effect sizes, while at the axes indicate sex-specific effects.

### 3.6 Sensitivity analyses

The summary of the results is shown in Table S14A-D. Although the Bonferroni significant CpGs varied across the sensitivity models, the effects sizes remained very similar to those of the main model with minor fluctuations (Figures S7-S11). The model adjusted for gestational age showed more similar effect sizes to the main model than the other sensitivity analyses (Figure S7). On the other hand, the analysis restricting to the participants with European origin presented the largest differences in effect sizes, however, their direction remained the same (Figure S8). Suggestive CpGs (p-value < 1E-05) of the sensitivity analyses are presented in Tables S15-S18.

### 3.7 Downstream analyses

The 27 Bonferroni-significant CpGs of the main model as well as the 74 sex-specific CpGs were annotated using different tools (Tables S4 for main model, S7 for sum of metabolites, S11 for sex-specific results). In the main model, we found that 25 out of the 27 CpGs were annotated to genes, 11 to CpG islands, and one (cg04915058 annotated to *C3orf51* and *ERC2* genes) had mQTLs. None of the CpGs was annotated to placental PMDs or gDMRs. In females and males, 33 and 30 of the CpGs were annotated to genes, eight and five to CpG islands, 17 and six to placental PMDs, and 10 and four had mQTLs. In males, one CpG (cg01224063 at gene *IGF1R*) was also annotated to a gDMR.

We, then, took suggestive CpGs from the main model (p-value <1E-05) and run pathway enrichment analyses for each phthalate and DINCH metabolite and period (going from 10 to 71 CpGs per analysis). At 19w, the identified pathways included immune and vascular regulation (MiBP), relaxin signaling (MiBP), signaling through growth factors (oh-MPHP), diabetes (MECPP) or regulation by androgen receptor (AR) (MEHHP) and estrogen receptor 1 (ESR1) (oxo-MINCH) (Table S19A), whereas at 35w, steroid biosynthesis (MECPP, sum DEHP) and anemia (MEHHP, MEOHP, sum DEHP), among others (Table S19B).

### 3.8 Comparison with previous studies

We compared our findings with those previously published in the EDEN and SEPAGES cohorts. In EDEN cohort, phthalates were measured in urine samples collected between 22w and 29w of pregnancy^35^. SEPAGES conducted both whole and sex-stratified analyses with whole pregnancy phthalate and DINCH exposures, defined as the median of levels measured at 18w and 34w of gestation^36^. None of our CpGs were among the list of FDR significant signals in any of the cohorts. FDR significant CpGs in EDEN and SEPAGES were not among suggestive CpGs in our results, regardless of the time period, the metabolites, or the main or sex-stratified analysis. A comparison of the FDR significant CpGs observed in cohorts EDEN and SEPAGES with our results is shown in Table S20. Most CpGs did not replicate, but some showed nominal associations in the same direction.

## 4. Discussion

Our study identified a number of associations between in utero exposure to phthalates and DINCH metabolites measured at two exposure windows (19w and 35w) and placental DNA methylation at birth. Most of the associations showed decreased DNA methylation with higher exposure levels.A substantial number of sex-specific associations were detected, which also differed across the exposure periods. Enrichment analysis revealed pathways important for placental function and fetal development.

### 4.1 Exposure window-specific effects

#### 4.1.1 Week 19 of gestation

At 19 weeks, we identified 21 unique CpGs annotated to 11 genes, six of which have been previously reported as differentially expressed following phthalate exposure in animal models according to CTD^65–69^. For example, *CROT* was downregulated in the livers of 12-week-old mice exposed to DEHP and altered in 72-hour-old frog embryos exposed to DMP^68,69^. In our study, 12 CpGs located in the promoter of *CROT* and the gene body of the lncRNA *TP53TG1* showed hypomethylation in association with MEP. *CROT* is involved in fatty acid oxidation, contributing to placental energy supply^70^. Another gene, *MIPEP*, was downregulated in fetal rat testes (gestational days 12–20) following DBP exposure^66^. In our study, *MIPEP* showed reduced methylation in the gene body in response to MEHP, and MEOHP. *MIPEP* is involved in oxidative phosphorylation, a critical energy-generating process in the placenta^71,72^. Notably, both *CROT* and *MIPEP* are highly expressed in cytotrophoblasts^73^. In addition, *FXYD2* was downregulated in bovine oocytes and blastocysts exposed to MEHP^67^. In our study, this gene showed lower DNA methylation at the gene body in association with MEHP exposure. *FXYD2* is part of the sodium:potassium-exchanging ATPase complex, important for cytotrophoblast function and nutrient transport during pregnancy^74^. We also observed increased promoter methylation of *ERC2* in association with higher levels of oh-MINCH and oxo-MINCH. *ERC2*, downregulated in fetal rat testes (gestational days 12–20) after DBP exposure^66^, has been linked to neurological and behavioral disorders in children^75,76^. Lastly, *RNPEP* was previously shown to be affected by DBP, DMP, and DCHP in fetal rat testes (gestational days 12–20) and in 72-hour-old frog embryos^66,69^. In our study, it showed lower methylation at the gene body with increasing cx-MiNP levels. *RNPEP* catalyzes arginine metabolism, essential for fetal development^77^.

Biological pathways in the placenta potentially altered by exposure to phthalates at 19w include signaling through growth factors, vascularization, and immune response. For example, the current study results highlight the influence of MiBP on the chemokine signaling pathway. Chemokines are important for accurate placental cell function and vascularization. Alterations in their expression may lead to pregnancy complications including preterm birth, preeclampsia or recurrent spontaneous abortion^78^. Furthermore, MiBP showed associations with Apelin and VEGF signaling and oh-MPHP was associated with FGF and PDFG signaling pathways. These pathways are involved in placental angiogenesis, besides VEGF and FGF are essential for its regular formation^79,80^). Several epidemiological and animal studies earlier reported vascular alterations in placenta related to phthalate exposure such as DEHP, DHP or DCHP^81^; however, none of them directly linked the disorders to MiBP or oh-MPHP. In addition, we observed alterations on relaxin signaling pathway related to MiBP. Relaxin ensures a relaxation of birth canal before delivery^82,83^.

We also investigated whether annotated genes were enriched for the regulation by certain transcription factors and found that MEHHP was associated with CpGs in genes regulated by the androgen receptor (AR). Previous *in vitro* and *in silico* studies have investigated the agonistic and antagonistic actions against AR of DEHP, the parental compound of MEHHP, and while no androgenic activity was observed in any of these studies, they reported conflicting results of its anti-androgenic activity^84–88^. Moreover, we also observed that oxo-MINCH was associated with CpGs annotated to genes regulated by the estrogen receptor 1 (ESR1). This is consistent with findings from an *in vitro* study, which reported ESR1 activation induced by DINCH metabolites, including oxo-MINCH^85^. In contrast, another *in vitro* study found no ESR1 activity from DINCH metabolites; however, this study used lower metabolite levels, which may account for the discrepancy^89^.

#### 4.1.2 Week 35 of gestation

At 35 weeks of gestation, six CpGs significantly associated with phthalate levels were annotated to five genes. Four of these genes have previously been reported as differentially expressed in phthalate-exposed animal models^66,67,90,91^. For example, *SLC36A4* was upregulated in the testes of adult male rats exposed to DEHP and downregulated in bovine oocytes and blastocysts exposed to MEHP, its primary metabolite^67,90^. In our study, CpGs within the *SLC36A4* gene body were hypermethylated in association with MECPP, MEHHP, and MEOHP. *SLC36A4* encodes an amino acid transporter involved in nutrient transfer across the placenta^92^. We also identified a differentially methylated CpG at the *RASSF1* promoter in association with MBzP exposure. *RASSF1* was previously shown to be upregulated in fetal testes of mice (gestational day 18) and rats (gestational days 12–20) following phthalate exposure^66,91^. This gene regulates cell cycle and apoptosis and has been linked to intrauterine growth restriction and preeclampsia^93,94^. In addition, CpGs in *ARMC8* (promoter) and *NDST1* (gene body) were differentially methylated in relation to MEHP. Both genes were downregulated in fetal testes of rats (gestational days 12–20) exposed to DBP^66^. *ARMC8* belongs to the armadillo superfamily involved in cellular communication, while *NDST1* plays a role in heparan sulfate biosynthesis and inflammation^95–97^. However, their roles in placental development have not been established.

The enrichment analyses at 35w highlighted, among other pathways, steroid biosynthesis (MECPP) and anemia (MEHHP and MEOHP). Whereas the association of anemia and phthalate exposure is not well documented, several studies have shown the impact of exposure to DEHP or MEHP on steroidogenesis in rat and mouse placentas^98–100^.

The different results observed between exposure windows could have a number of explanations. It could be that the exposure to phthalate and DINCH metabolite might affect the expression of different genes/pathways at each period, which we are able to observe at birth through permanent and distinct DNA methylation marks. According to this hypothesis, it would seem that early in pregnancy (19w) phthalates and DINCH affect placental formation (i.e. vascularization, growth factors) and the regulation of genes that respond to estrogen or androgen hormones; while at later stages (35w) they influence the production of steroid hormones. It could also be that the incomplete overlap of mother-child pairs at each time point have influenced the results leading to different lists of Bonferroni-adjusted significant CpGs at each exposure period.

### 4.2 Sex-specific effects

We identified a substantial number of differentially methylated CpGs associated with phthalate and DINCH metabolites in a sex-specific manner at 19w and 35w. This goes in line with the expected sex-specific endocrine disruptor activity of the phthalate and DINCH metabolites^101^, and with the sex-specific epigenetic pattern of the placenta^102,103^.

Female-specific results include associations between MiBP levels and cg00752628 annotated to the *RSPO3* gene, and between cx-MiNP levels and cg22505312, cg22505312 and cg23710890 annotated to the *CTSB* gene at 19w. Whereas *RSPO3* has not been previously associated with exposure to phthalates in animal and *in vitro* studies, *CTSB* has been found overexpressed in frog embryos exposed to DCHP, DMP or MMP^69^.

Among male-specific associations, we identified higher DNA methylation at four CpGs of the *IGF1R* gene body in relation to higher levels of oh-MPHP at 35w. One of the CpGs (cg01224063) was located in a maternal gDMR described in the placenta^56^. The insulin□like growth factors are involved in placental and fetal growth, carrying out various functions including trophoblast proliferation, migration, and invasion^104^. Another interesting male-specific association was observed between levels of oxo-MINCH at 19w and lower DNA methylation at cg23939844 annotated to *LRTM2* and *CACNA2D4* genes. *CACNA2D4* has been previously reported to be underexpressed in human embryonic stem cells in relation to exposure to DEHP^105^.

### 4.3 Mixture analyses

The mixture analysis identified six significant associations of the overall 27 tested CpGs. Although in the analysis of individual compounds the metabolites showed mostly the effects in the same direction on the individual CpGs, in the mixture analyses, the partial effects were opposite. For instance, at 35w, MEHHP, MEOHP, MECPP were individually significantly associated with higher DNA methylation of cg07964716. However, in the mixture analysis MEHHP showed a negative effect on this CpG. This can be explained by the high correlation among these metabolites, which could lead to reversed partial effects in the mixture model. Only a few studies have performed mixture analyses relating combined exposure to phthalates and phenols with cord blood DNA methylation^29,33^, but none have identified CpGs overlapping with those found in our study.

### 4.4 Comparison with previous studies

The lack of replication of the CpGs observed in cohorts EDEN and SEPAGES could have been resulted due various reasons. First, the exposure assessment was not equivalent in the three cohorts in relation to the exposure window (22-29w in EDEN, whole pregnancy in SEPAGES and 19w and 35w in BiSC), number of urine voids tested (1 in EDEN, and >10 in the other two cohorts) or the laboratory (EDEN: National Center for Environmental Health; BiSC, SEPAGES: Norwegian Institute of Public Health). While EDEN used the array Infinium HumanMethylation450 BeadChip, BiSC and SEPAGES used the array Infinium Methylation EPIC BeadChip. Additionally, the population characteristics (ie. ethnicity, socio-economic status, etc.), exposure levels, and statistical modelling may have potentially influenced the results.

### 4.5 Strengths and limitations

Our study had several strengths. First, exposure assessment involved the first and last voids of the day over six consecutive days (10-12 samples per urine pool) ensuring high robustness and accuracy of our exposure assessment compared to studies based on a single urine void. Second, we analyzed data from two exposure windows, which allows a comprehensive assessment of phthalate exposure across different stages of placental and fetal development. Moreover, with sample sizes ranging from 349 to 404, this study modestly exceeds the size of previous research. Third, the EWAS was performed using robust linear models able to control for outliers, providing more reliable estimates. Moreover, by conducting sensitivity analyses, we showed the robustness of our observed findings. Last, we conducted mixture analyses providing deeper and more realistic insights into the impact of whole exposure on placental DNA methylation in comparison to analyses of individual chemicals.

However, this study also faced some limitations. First, although we used the EPIC array, which contains >0.8M CpG sites, we still did not cover all possible CpG sites across the genome, limiting our ability to detect all potential DNA methylation changes associated with the exposures and outcomes of interest. Second, given that our analyses were based on bulk tissue, we could not assess the cell type-specific effects, potentially overlooking the distinct roles that different cell types may play. Finally, although we considered a wide range of relevant covariates in our analyses, residual confounding cannot be completely ruled out, and causal inference requires of complementary studies such as interventional studies or *in vitro* or *in vivo* models.

## 5. Conclusion

Our study suggests that in utero exposure to phthalate and DINCH metabolites is associated with differential placental DNA methylation in pathways related to steroid synthesis and action, vascularization and immunity. These epigenetic modifications were specific to the timing of exposure. Moreover, sex-specific effects were observed, consistent with the known endocrine-disrupting properties of phthalate and DINCH metabolites. Further studies are needed to confirm these findings and to determine whether the placental epigenetic signatures linked to in utero exposure have implications for short- and long-term health effects.

## Supporting information

List of Supplementary Figures

List of Supplementary Tables

## Data Availability

Genome-wide DNA methylation summarized results will be uploaded to Zenodo repository (https://doi.org/10.5281/zenodo.15746984) and will be available after publication of the paper. Individual level data may be available by contating BiSC (https://projectebisc.org/en/contact/). Code can be shared upon request with the authors.

## Declaration of competing interest

The authors declare that they have no known competing financial interests or personal relationships that could have appeared to influence the work reported in this paper.

## Funding

This study was funded by the European Union’s Horizon 2020 research and innovation programme-EU.3.1.2. (874583 - ATHLETE project). The BiSC cohort has received funding from the European Research Council (ERC) under the European Union’s Horizon 2020 research and innovation programme (785994 – AirNB project), from the AGAUR-Agència de Gestió d’Ajuts Universitaris de Recerca (2017 SGR 826 - Population Neuroscience group), from the Centro de Investigación Biomédica en Red de Epidemiología y Salud Pública (CIBERESP) (CB06/02/0041), from the Instituto de Salud Carlos III (ISCIII) and the European Regional Development Fund (ERDF) - Maternal and Child Health and Development Network (SAMID) (RD16/0022/0014 and RD16/0022/0015), and from the Instituto de Salud Carlos III (ISCIII) and the European Union Next Generation EU - Primary Care Interventions to Prevent Maternal and Child Chronic Diseases of Perinatal and Developmental Origin Network (RICORS-SAMID) (RD21/0012/0001 and RD21/0012/0003). Placental DNA methylation was funded by the Instituto de Salud Carlos III (ISCIII) and co-funded by European Union (ERDF) “A way to make Europe” (PI20/00190 – ALMA project), and from the European Joint Programming Initiative “A Healthy Diet for a Healthy Life” (JPI HDHL and Instituto de Salud Carlos III) (AC18/00006 - NutriPROGRAM project). ISGlobal acknowledges support from the grant CEX2023-0001290-S funded by MCIN/AEI/10.13039/501100011033, and support from the Generalitat de Catalunya through the CERCA Program.

## Acknowledgements

We would like to thank all the participants and their families for their generous collaboration. A full list of BiSC researchers can be found at https://projectebisc.org/en/team/.

## CRediT authorship contribution statement

**Hana Vespalcova:** Data curation, Conceptualization, Methodology, Software, Formal analysis, Data curation, Writing - Original Draft, Writing - Review & Editing, Visualization. **Bethany Knox:** Data curation, Writing - Review & Editing. **Amrit Kaur Sakhi:** Resources, Methodology, Writing - Review & Editing. **Cathrine Thomsen:** Resources, Methodology, Writing - Review & Editing. **Sofía Aguilar-Lacasaña:** Data curation, Writing - Review & Editing. **Marta Cosin-Tomas:** Data curation, Writing - Review & Editing. **Laura Gómez-Herrera:** Data curation, Writing - Review & Editing. **Olga Sánchez García:** Resources, Writing - Review & Editing. **Elisa Llurba:** Resources, Writing - Review & Editing. **María Dolores Gómez-Roig:** Resources, Writing - Review & Editing. **Jordi Sunyer:** Resources, Funding acquisition,Writing - Review & Editing. **Payam Dadvand:** Resources, Funding acquisition,Writing - Review & Editing. **Mariona Bustamante:** Conceptualization, Methodology, Writing - Review & Editing, Supervision. **Martine Vrijheid:** Conceptualization, Methodology, Funding acquisition,Writing - Review & Editing, Supervision.

## Ethics

The BiSC study, from pregnancy up to the 18 month-visit, was approved by the Clinical Research Ethics Committee of the Parc de Salut Mar project (2018/8050/I), Medical Research Committee of the Fundació de Gestió Sanitària del Hospital de la Santa Creu i Sant Pau de Barcelona (EC/18/206/5272), and Ethics Committee of the Fundació Sant Joan de Déu (PIC-27-18). Before joining the cohort during their regular first trimester hospital visit, participants were informed by a BiSC midwife or nurse about the study’s details, duration, and their option to withdraw without penalty. If they agreed to take part, they signed consent forms permitting the collection of biological samples and genetic studies, receiving a copy for themselves.

## Notes

### Competing Interest Statement

The authors have declared no competing interest.

### Author Declarations

Clinical Research Ethics Committee of the Parc de Salut Mar project (2018/8050/I), Medical Research Committee of the Fundacio de Gestio Sanitaria del Hospital de la Santa Creu i Sant Pau de Barcelona (EC/18/206/5272), Ethics Committee of the Fundacio Sant Joan de Deu (PIC-27-18).

