## Supplementary figures and images for "Association of in utero exposure to phthalate and DINCH metabolites with placental DNA methylation"

### Supplementary Figure 1

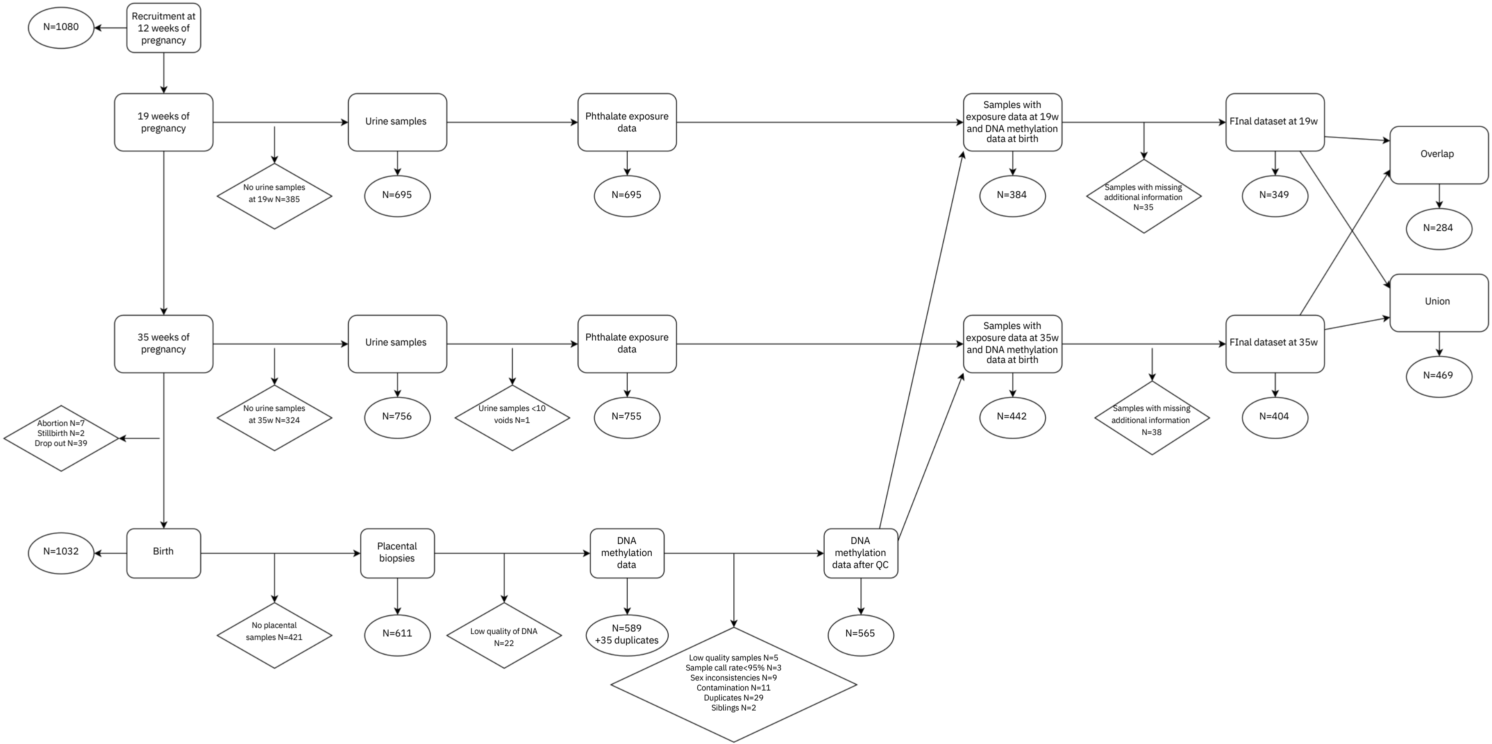

### Supplementary Figure 2

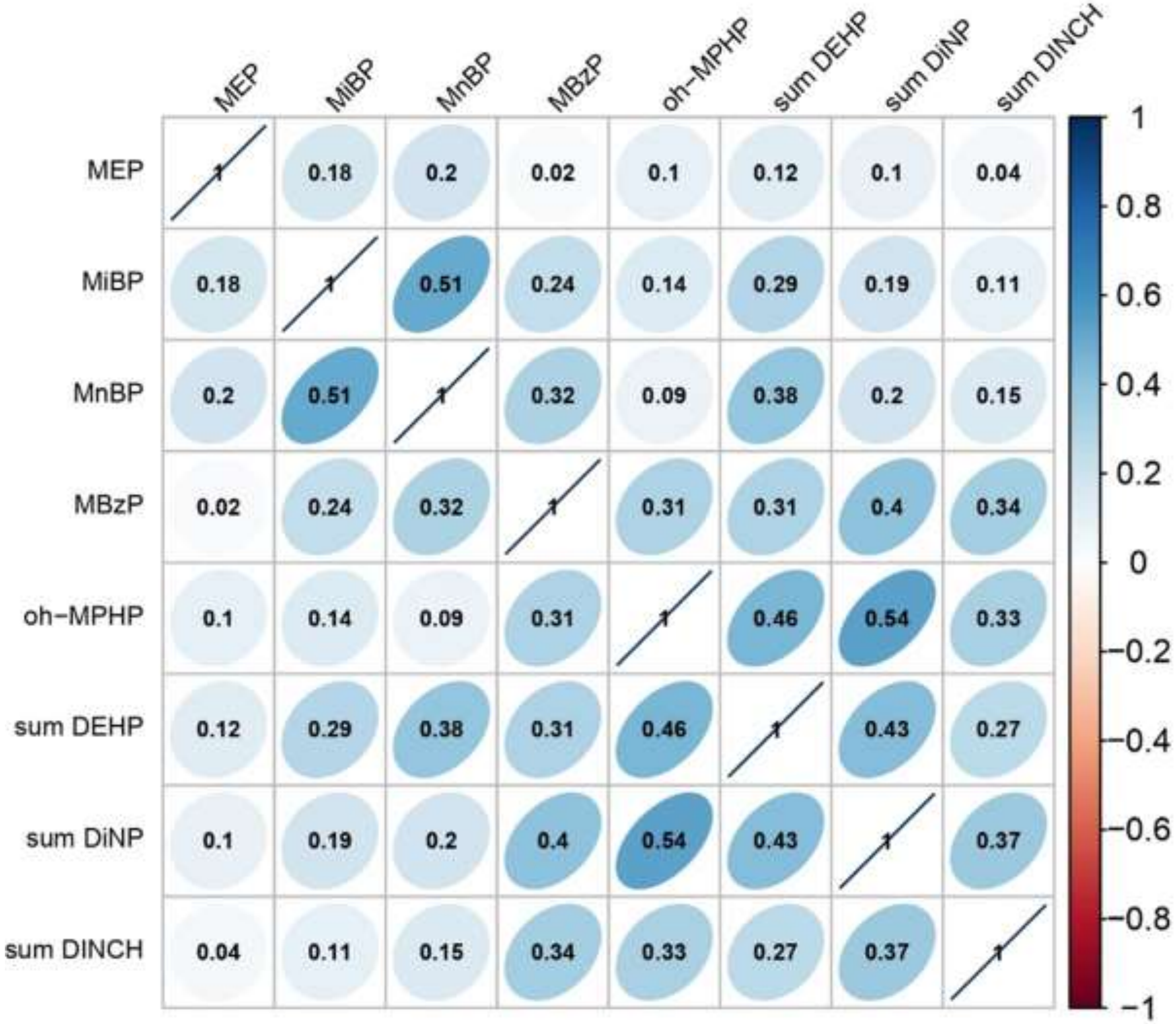

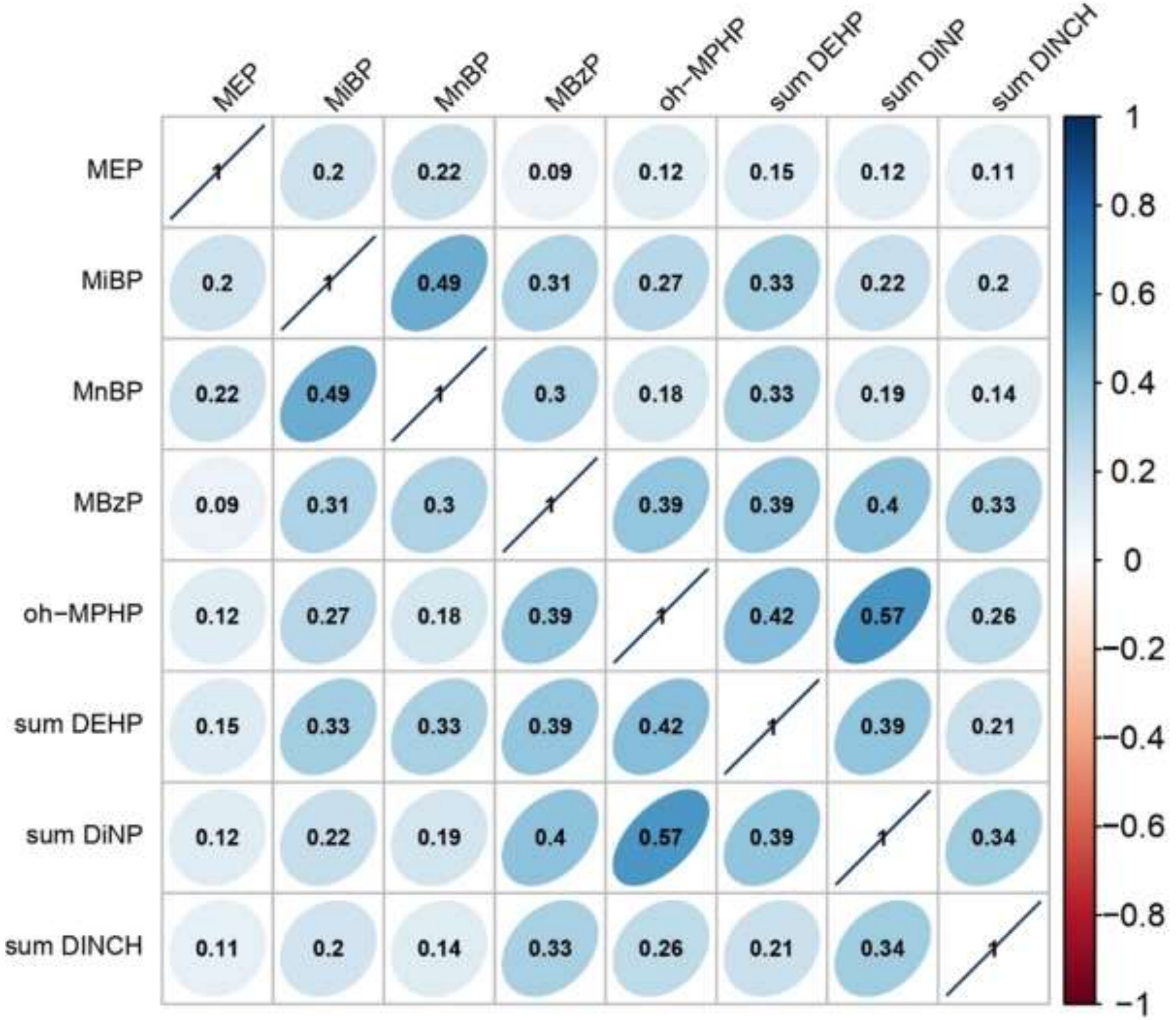

### Supplementary Figure 3

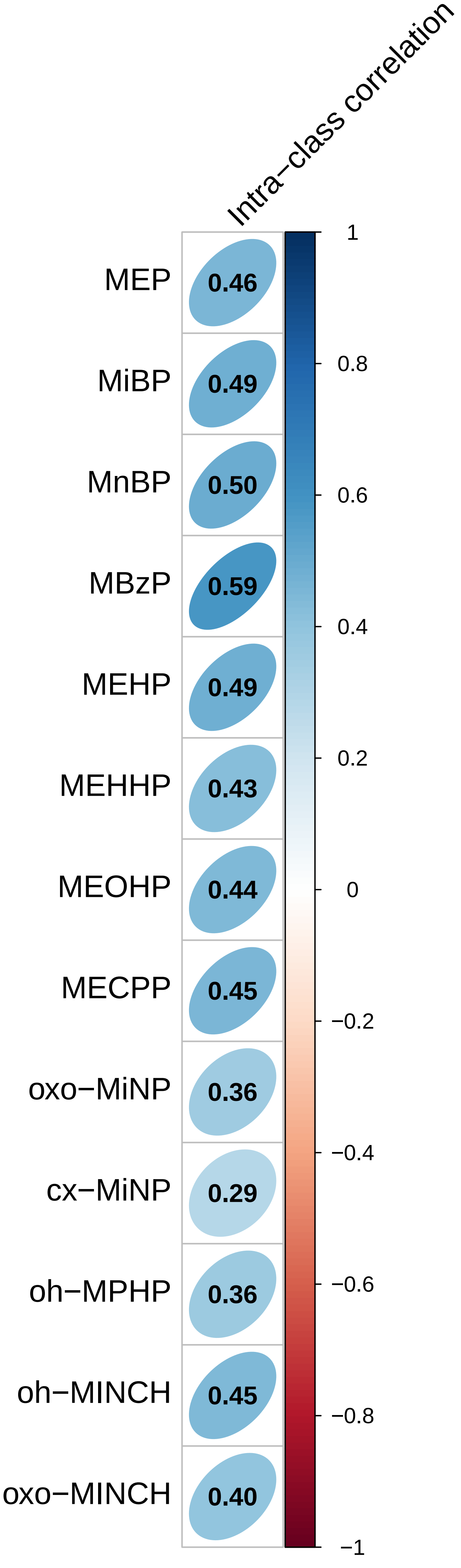

### Supplementary Figure 4

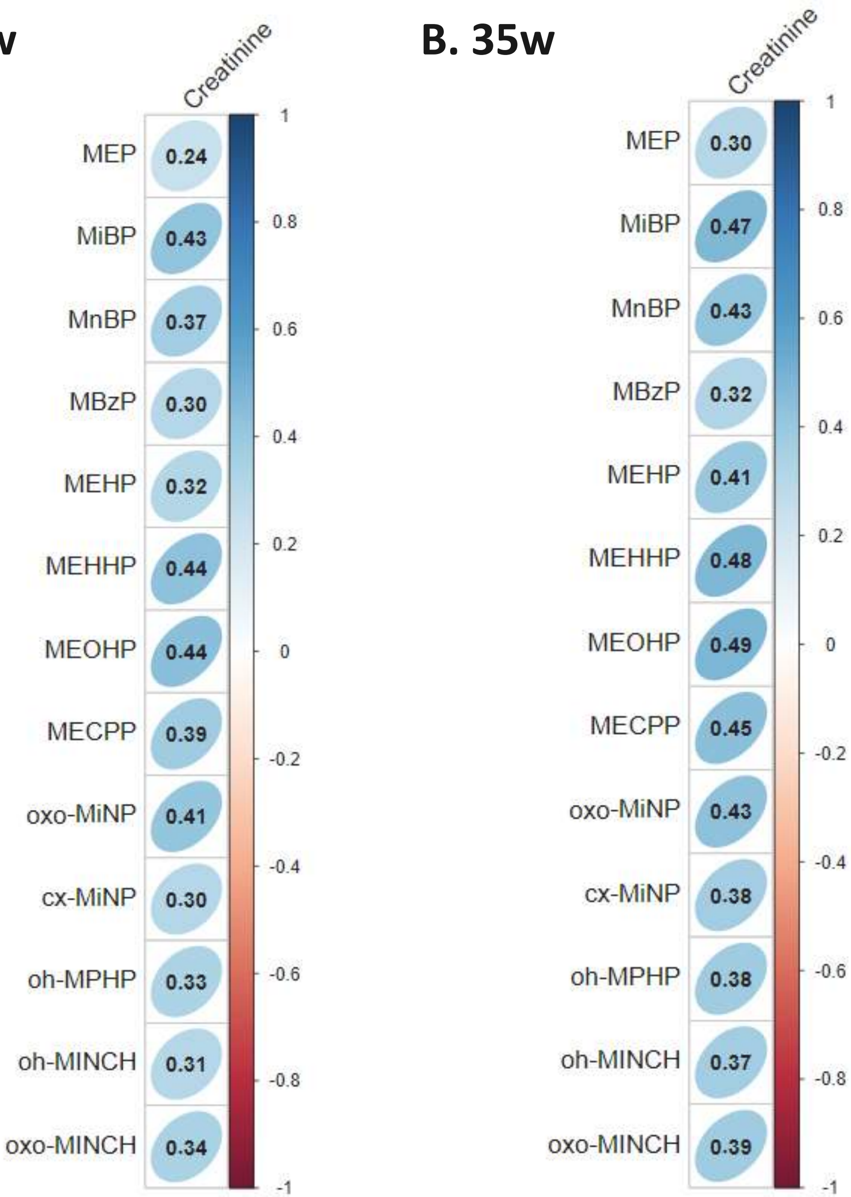

### Supplementary Figure 5A

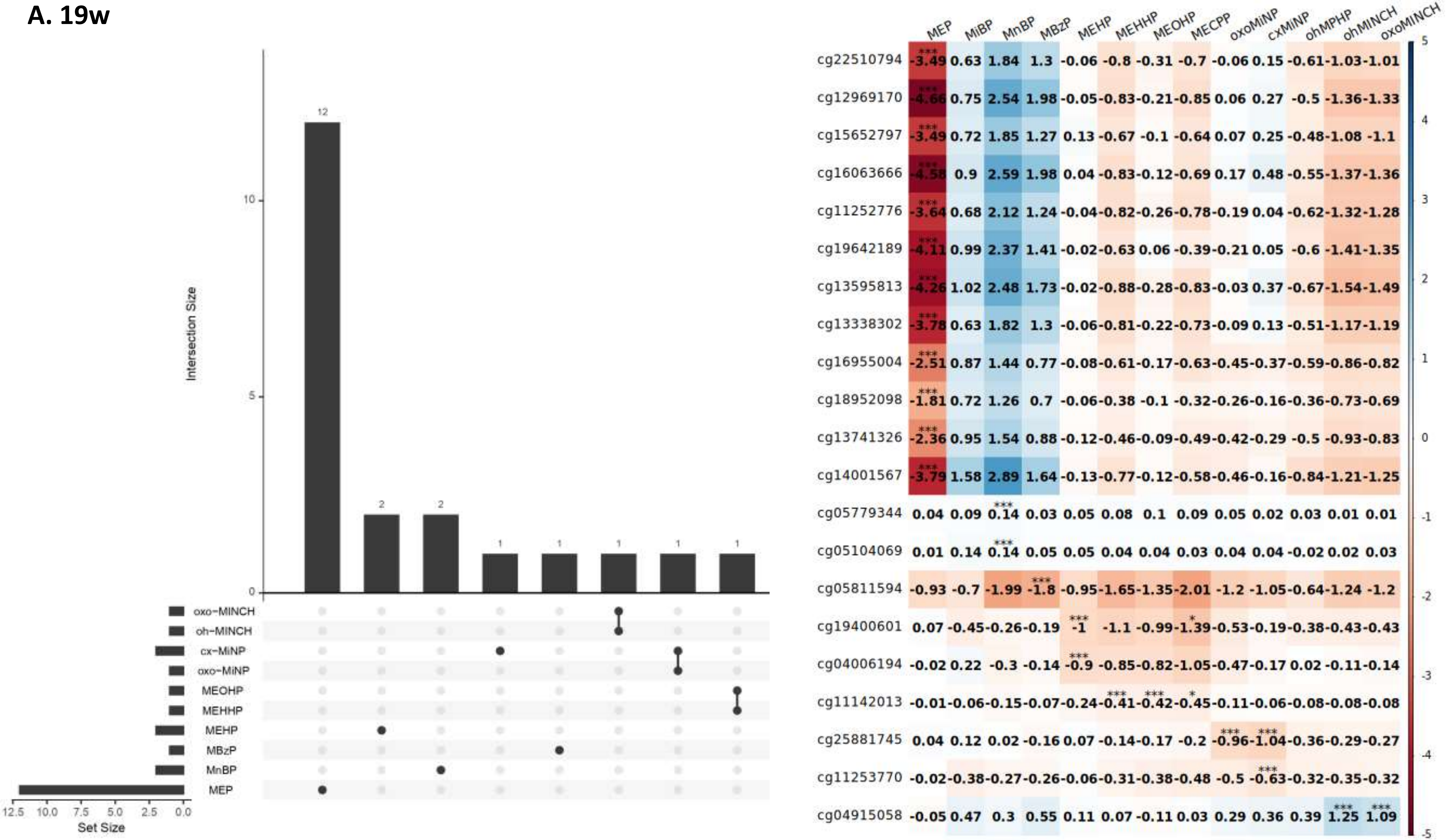

### Supplementary Figure 5B

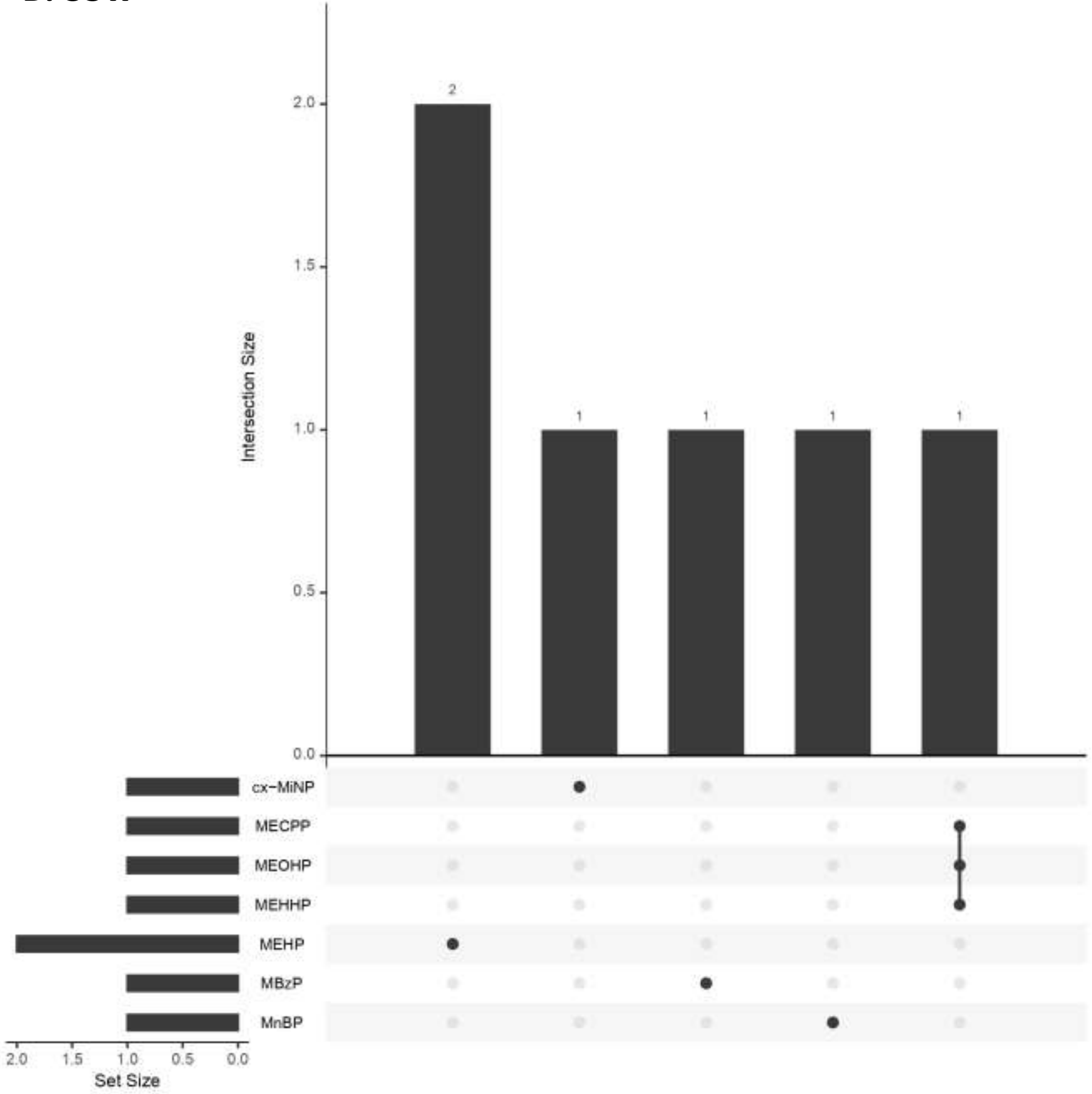

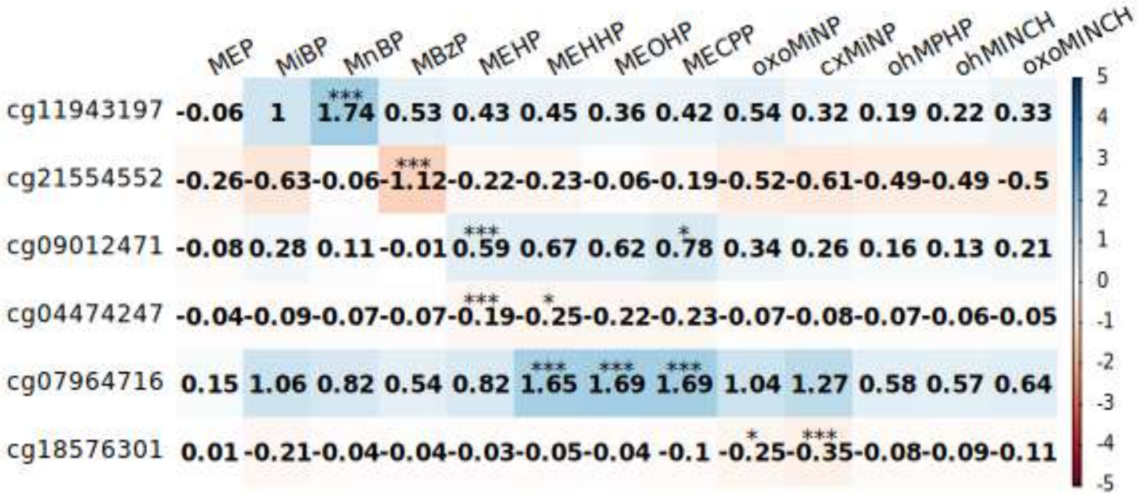

### Supplementary Figure 6

#### A. Partial effect sizes on significant CpGs at 19w

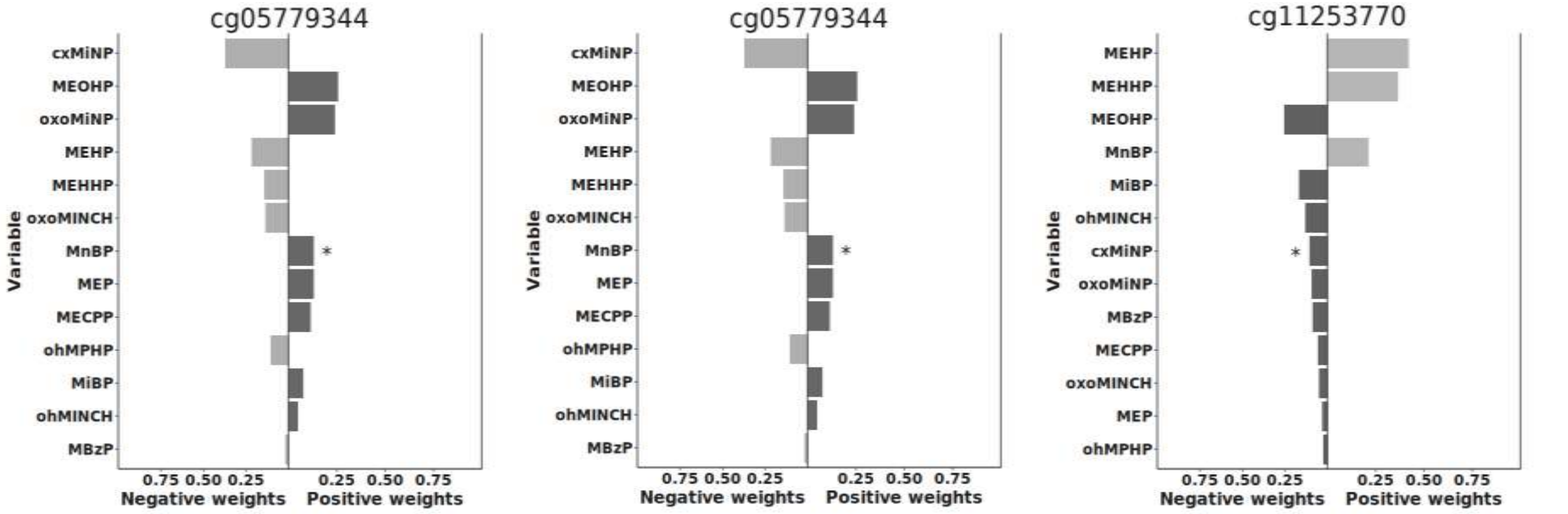


#### B. Partial effect sizes on significant CpGs at 35w

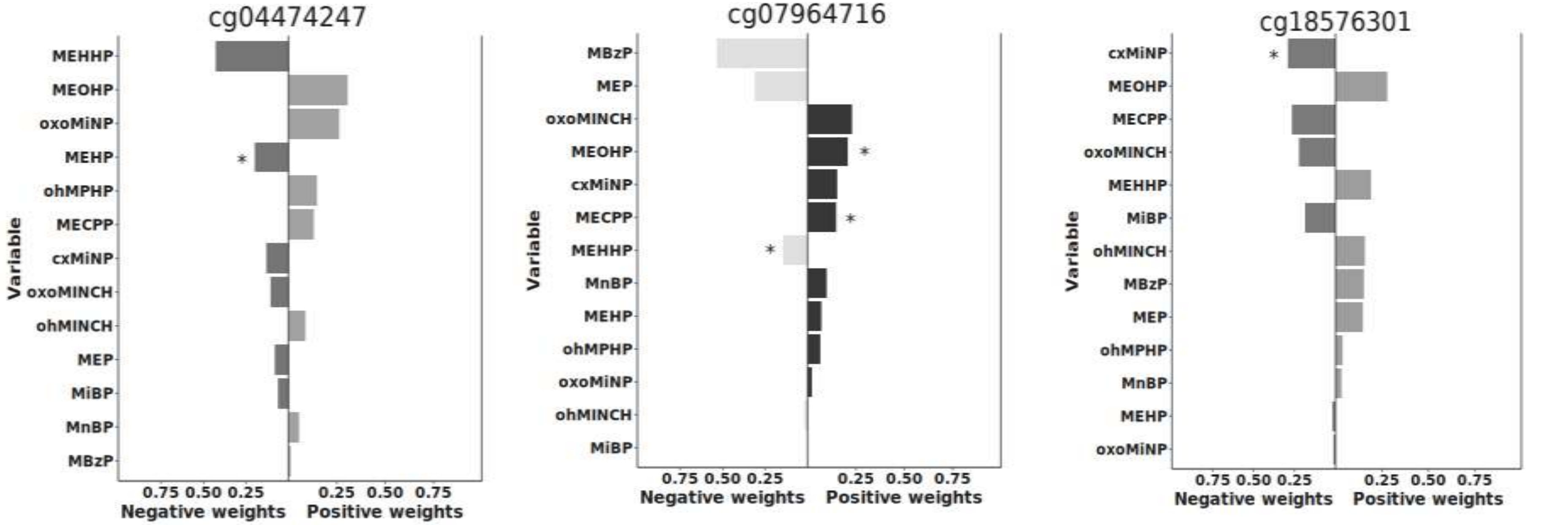

### Supplementary Figure 7

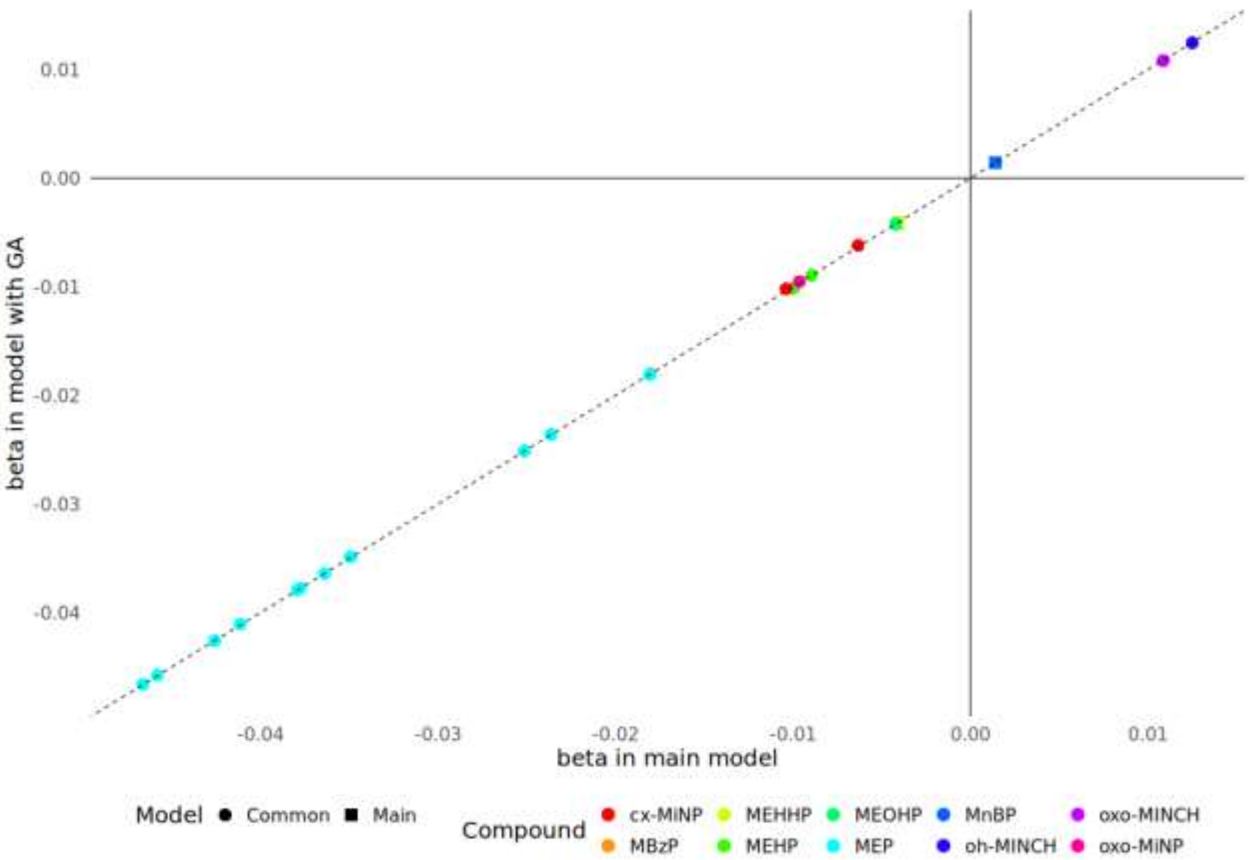

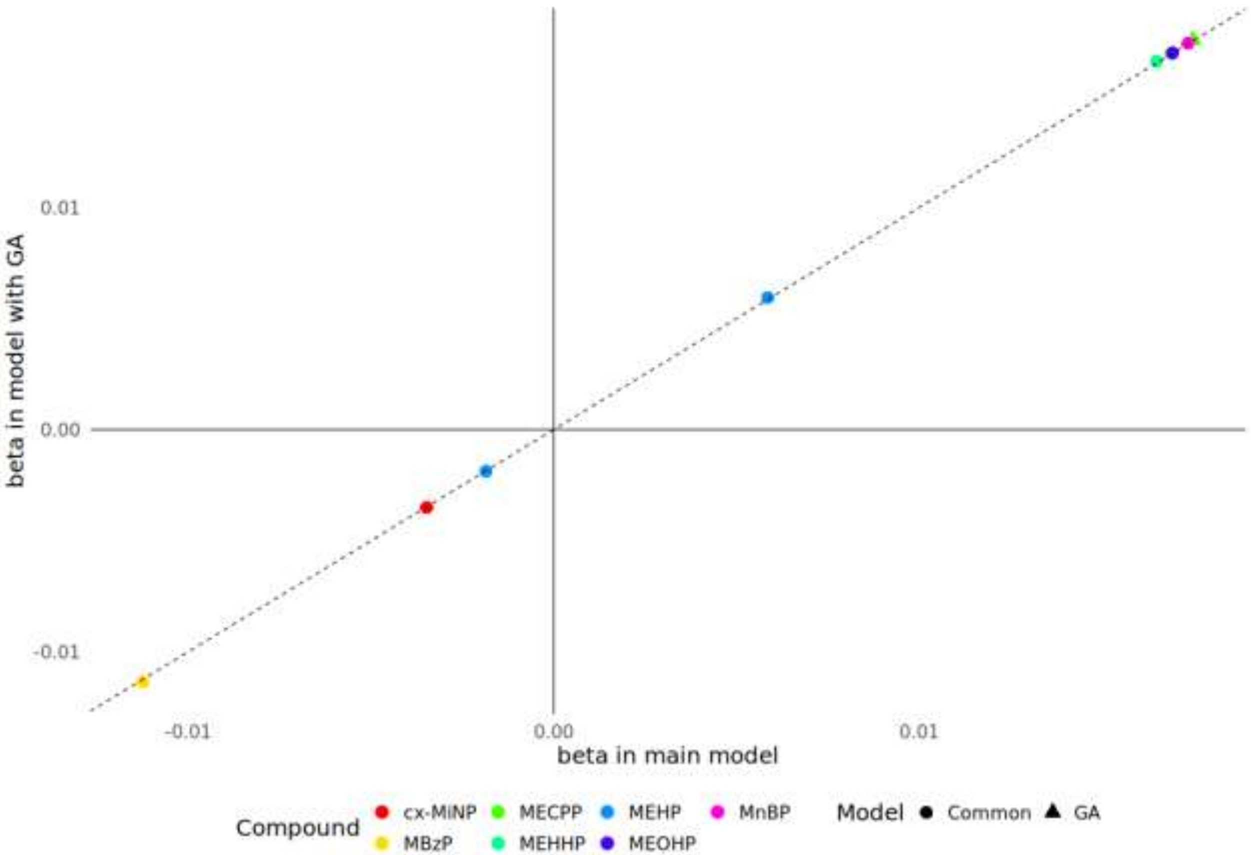

### Supplementary Figure 8

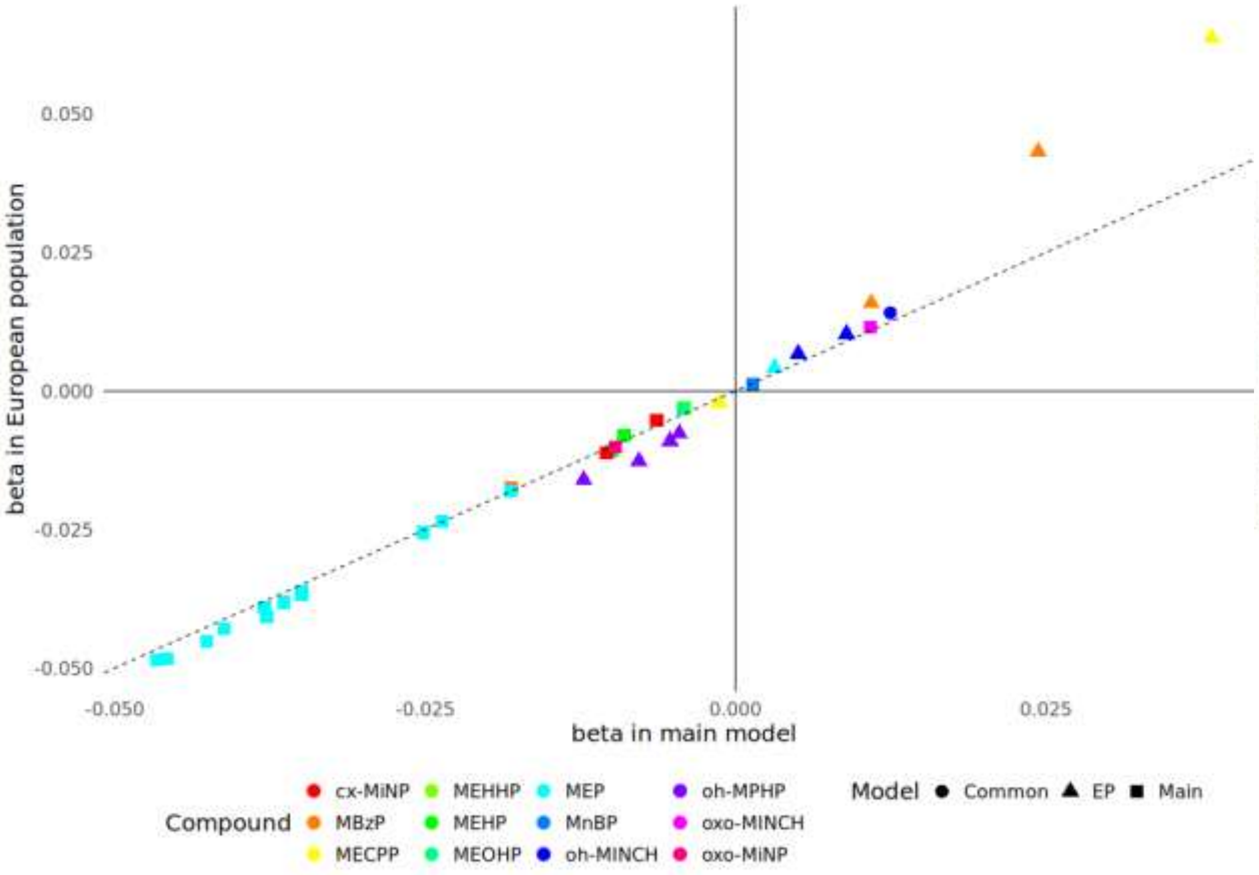

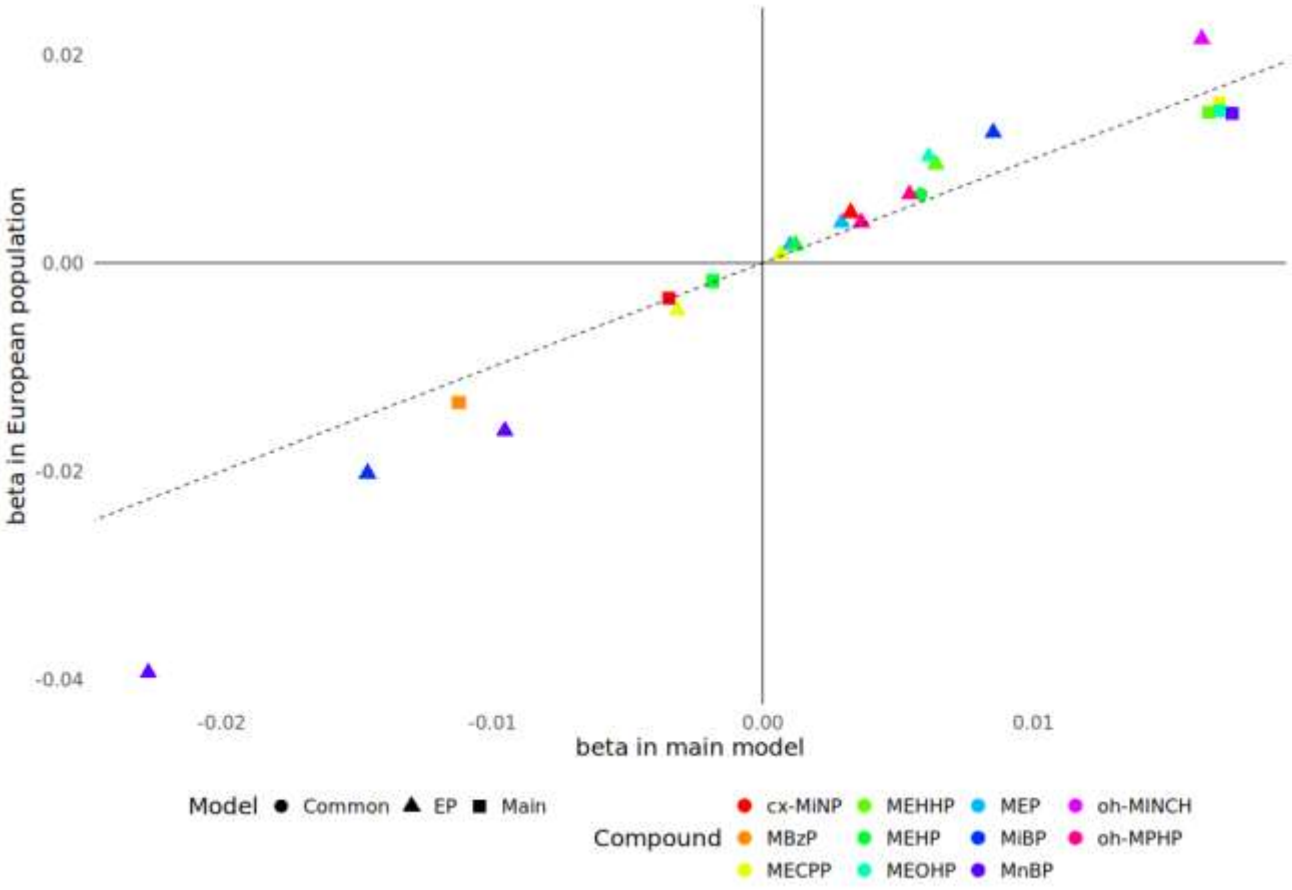

### Supplementary Figure 9

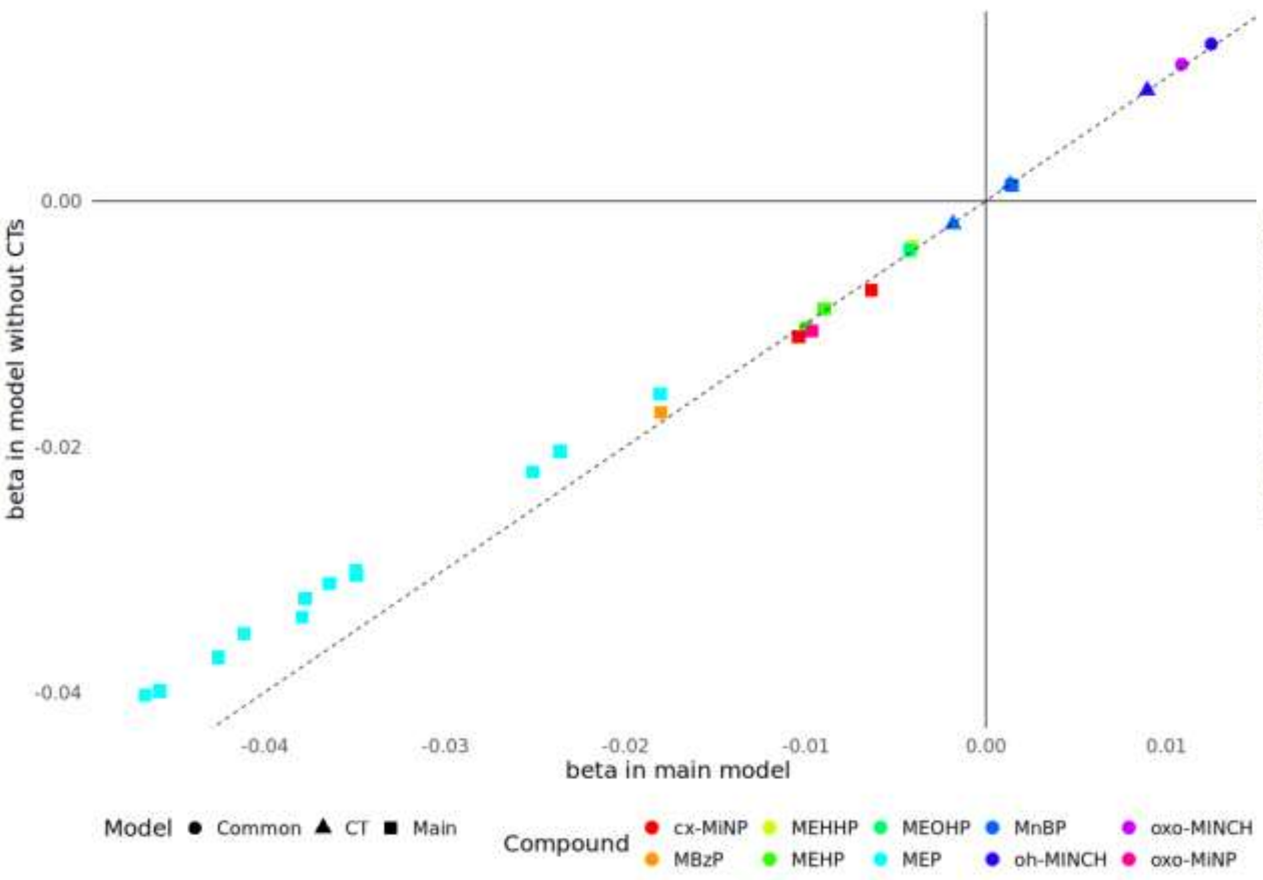

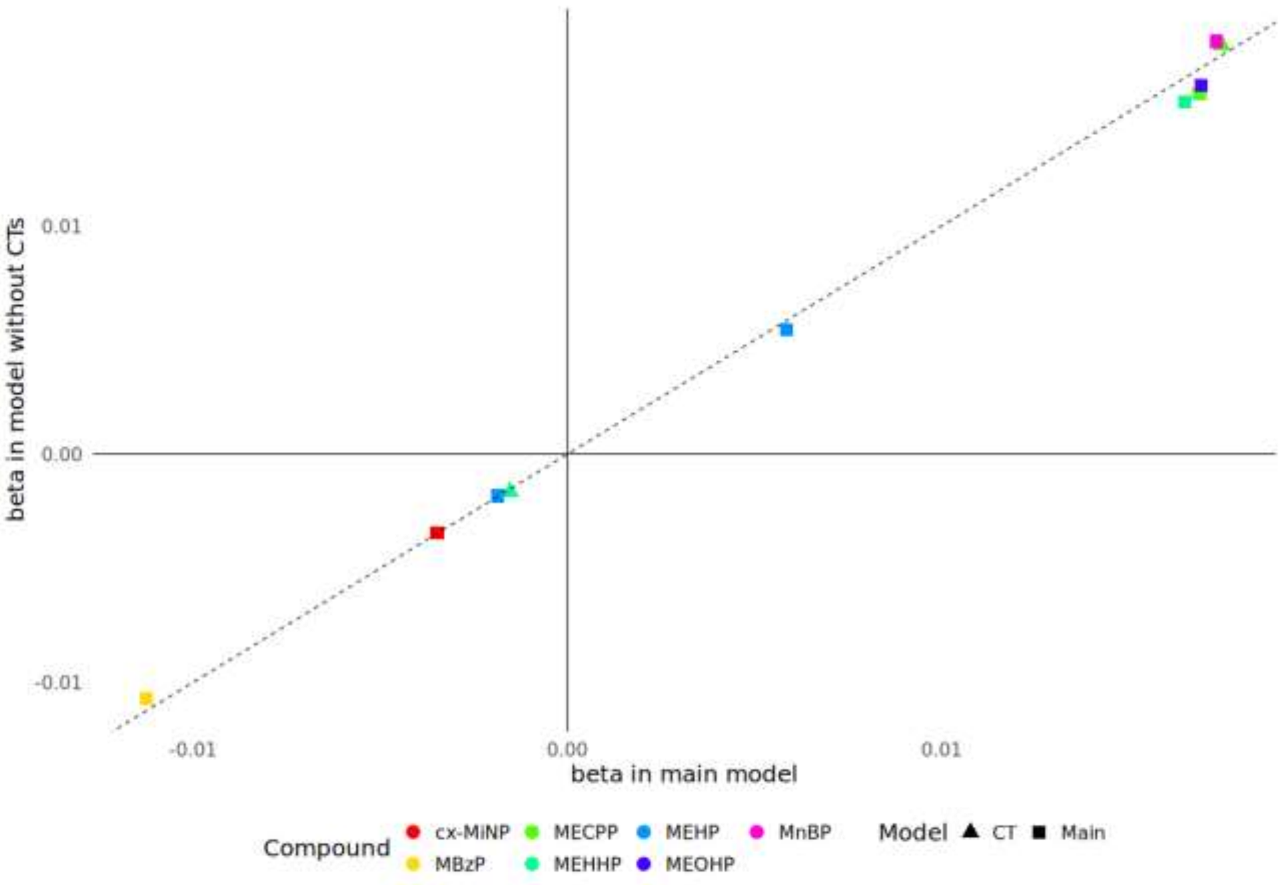

### Supplementary Figure 10

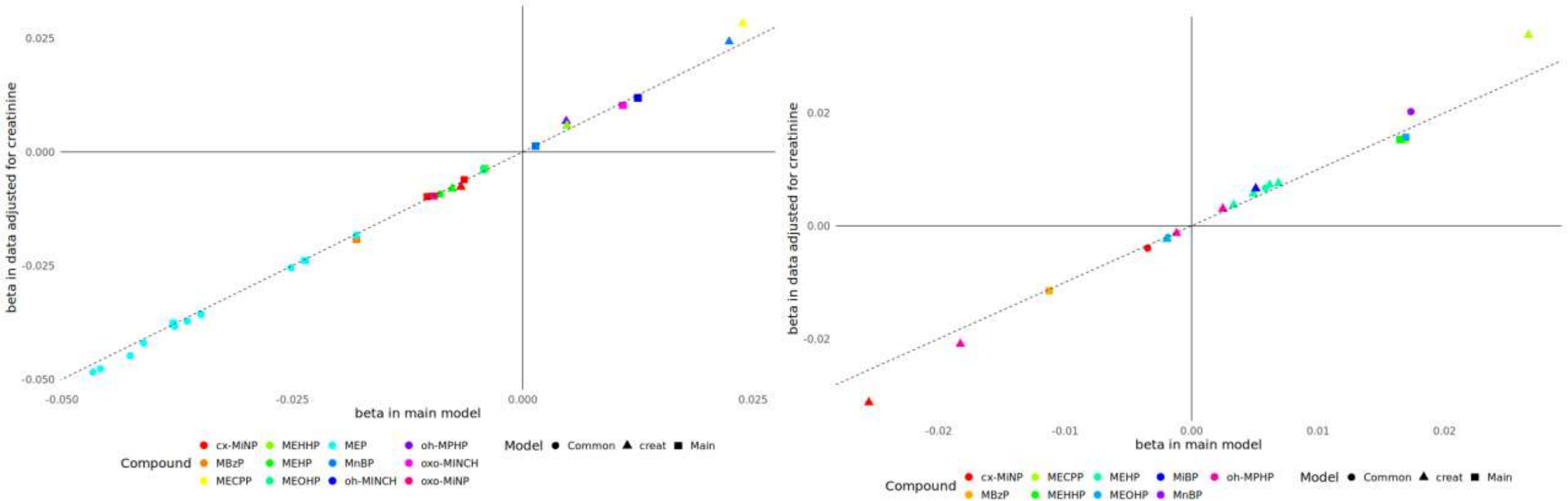

### Supplementary Figure 11

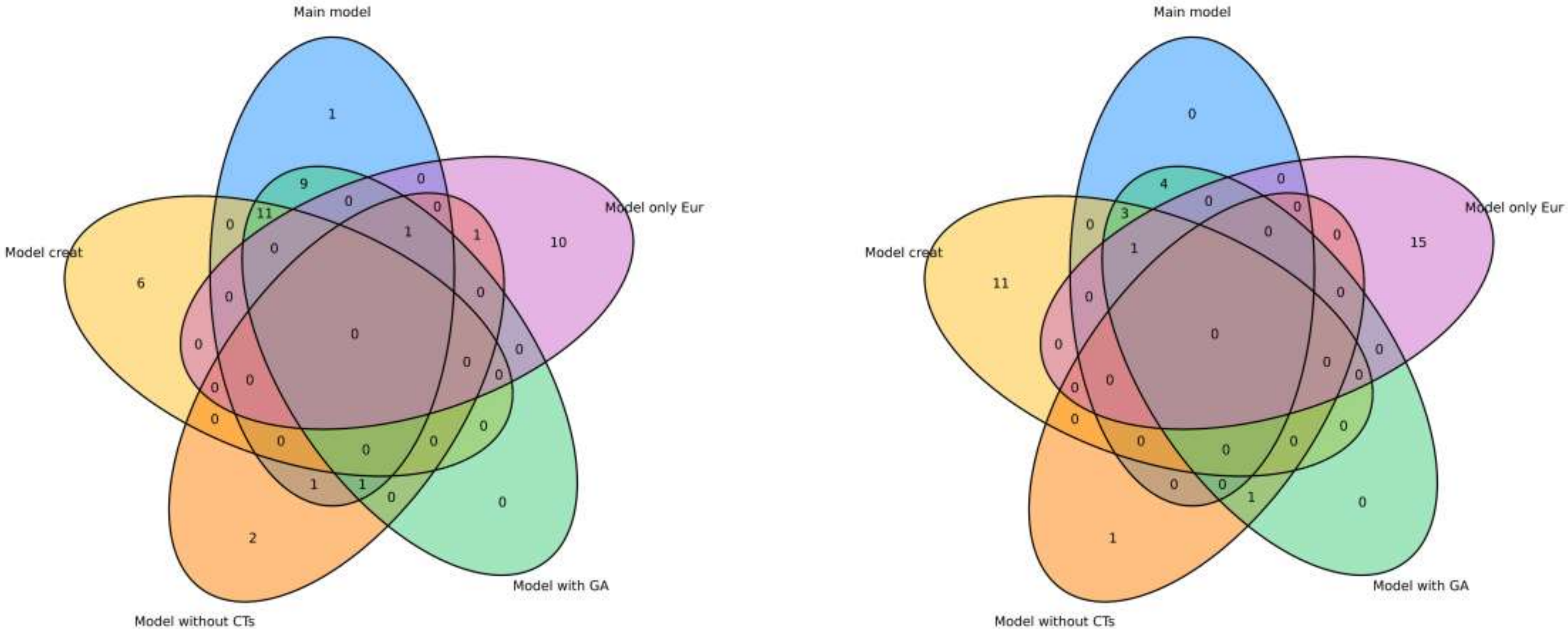
