## Supplementary material for "Association of in utero exposure to phthalate and DINCH metabolites with placental DNA methylation": List of Supplementary Figures

**Figure S1. Number of participants in the study.**

The flowchart shows the number of participants in the Barcelona Life Study Cohort (BiSC), exclusion criteria and final number of participants included in this study.

**Figure S2A. Correlation of phthalate and DINCH metabolite levels, by exposure period.**

The heatmap shows the Pearson’s correlation of the levels of phthalate and DINCH metabolites, and the sums measured at 19w (A) and 35w (B). Positive correlations are indicated in blue, and inverse correlations in red.

A. 19w
B. 35w

**Figure S3.** **Intra-class correlation of phthalates and DINCH metabolites between pregnancy exposure periods.**

The heatmap shows the intra-class and inter-class correlation of the levels of 13 phthalate and DINCH metabolites. Positive correlations are indicated in blue, and inverse correlations in red.

**Figure S4. Correlation of phthalate and DINCH metabolites with creatinine and specific gravity, by exposure period.**

The heatmap shows the Spearman’s correlation of the levels of 13 phthalate and DINCH metabolites with creatinine measured at 19w (A) and 35w (B). Positive correlations are indicated in blue, and inverse correlations in red.

A. 19w
B. 35w

**Figure S5. Overlap and correlation of effect sizes across phthalate and DINCH metabolites, by exposure period.**

Upset plot showing the overlap of Bonferroni significant CpGs across phthalate and DINCH metabolites measured at 19w (A) and at 35w (B). Heatmap plot showing the effect sizes of the association with DNA methylation across phthalate and DINCH metabolites measured at 19w (A) and at 35w (B). Suggestive associations (p value <1E-05) are indicated by one asterisk, and associations passing Bonferroni multiple-testing (p value <1e-07) by three asterisks.

A. 19w
B. 35w

**Figure S6. Partial effect sizes of the association of DNA methylation and phthalate and DINCH metabolites derived from the mixture analyses**.

The significant CpG-metabolite pair associations are indicated by asterix.

A. Partial effect sizes on significant CpGs at 19w of pregnancy
B. Partial effect sizes on significant CpGs at 35w of pregnancy

**Figure S7.** **Correlation of effect sizes between main model and the sensitivity model adjusted for gestational age, by exposure period.**

Each symbol represents the association of the DNA methylation levels at one CpG with the levels of one of the phthalate or DINCH metabolites measured at 19w (A) and at 35w (B). The x-axis represents the effect size in the main model, and the y-axis in the sensitivity model. The shape of the symbols represent the model and the color the phthalate or DINCH metabolite (see legend). Symbols at the diagonal indicate similar effect sizes, while at the axes indicate period specific effects.

A. 19w
B. 35w

**Figure S8. Correlation of effect sizes between the main model and the sensitivity model restricted to children of European origin, by exposure period.**

Each symbol represents the association of the DNA methylation levels at one CpG with the levels of one of the phthalate or DINCH metabolites measured at 19w (A) and at 35w (B). The x-axis represents the effect size in the main model, and the y-axis in the sensitivity model. The shape of the symbols represent the model and the color the phthalate or DINCH metabolite (see legend). Symbols at the diagonal indicate similar effect sizes, while at the axes indicate period specific effects.

A. 19w
B. 35w

**Figure S9. Correlation of effect sizes between the main model and the sensitivity model unadjusted for cellular composition, by exposure period.**

Each symbol represents the association of the DNA methylation levels at one CpG with the levels of one of the phthalate or DINCH metabolites measured at 19w (A) and at 35w (B). The x-axis represents the effect size in the main model, and the y-axis in the sensitivity model. The shape of the symbols represent the model and the color the phthalate or DINCH metabolite (see legend). Symbols at the diagonal indicate similar effect sizes, while at the axes indicate period specific effects.

A. 19w
B. 35w

**Figure S10. Correlation of effect sizes between the main model and the sensitivity model adjusted for creatinine levels, by exposure period.**

Each symbol represents the association between the DNA methylation levels at one CpG and one of the phthalates or DINCH metabolites, at 19w (A) and at 35w (B). The x-axis represents the effect size in the main model, and the y-axis in the sensitivity model. The shape of the symbols represent the model and the phthalate or DINCH metabolite (see legend). Symbols at the diagonal indicate similar effect sizes, while at the axes indicate period specific effects.

A. 19w
B. 35w

**Figure S11. Overlap of significant associations between the main and sensitivity models, by exposure period.**

A. 19w
B. 35w
