## Supplementary material for "Association of in utero exposure to phthalate and DINCH metabolites with placental DNA methylation": List of Supplementary Tables

**Table S1.**

Table S1A. Detection and quantification levels of the phthalate and DINCH metabolites in maternal urine samples collected at 19w (N=349)

Table S1B. Detection and quantification levels of the phthalate and DINCH metabolites in maternal urine samples collected at 35w (N=404)

**Table S2.**

Table S2A. Descriptives of the phthalate and DINCH metabolite levels in maternal urine samples collected at 19w (N=349)

Table S2B. Descriptives of the phthalate and DINCH metabolite levels in maternal urine samples collected at 35w (N=404)

**Table S3. Summary of results of the main model at 19w and 35w**

**Table S4. Association of placental DNA methylation with phthalate and DINCH metabolite levels measured at 19w and 35w (only significant Bonferroni CpGs ordered by position)**

**Table S5.**

Table S5A. Association of placental DNA methylation with phthalate and DINCH metabolite levels measured at 19w (only suggestive significant (p-value<1E-05) CpGs ordered by position)

Table S5B. Association of placental DNA methylation with phthalate and DINCH metabolite levels measured at 35w (only suggestive significant (p-value<1E-05) CpGs ordered by position)

**Table S6. Summary of results of the main model at 19w and 35w, for the sum of metabolites**

**Table S7. Association of placental DNA methylation with sum of metabolites measured at 19w and 35w (only significant Bonferroni CpGs ordered by position)**

**Table S8.**

Table S8A. Association of placental DNA methylation with sum of metabolites measured at 19w with CpGs (only suggestive significant (p-value<1E-05) CpGs ordered by position)

Table S8B. Association of placental DNA methylation with sum of metabolites measured at 35w with CpGs (only suggestive significant (p-value<1E-05) CpGs ordered by position)

**Table S9.**

Table S9A. Summary of the mixture analyses at 19w

Table S9B. Summary of the mixture analyses at 35w

**Table S10.**

Table S10A. Summary of results of the main model in males, at 19w and 35w

Table S10A. Summary of results of the main model in females, at 19w and 35w

**Table S11. Association of placental DNA methylation with phthalate and DINCH metabolite levels measured at 19w and 35w by sex (only significant Bonferroni CpGs ordered by position)**

**TableS12.**

Table S12A. Association of placental DNA methylation with phthalate and DINCH metabolite levels measured at 19w in males (only suggestive significant (p-value<1E-05) CpGs ordered by position)

Table S12B. Association of placental DNA methylation with phthalate and DINCH metabolite levels measured at 35w in males (only suggestive significant (p-value<1E-05) CpGs ordered by position)

**TableS13.**

Table S13A. Association of placental DNA methylation with phthalate and DINCH metabolite levels measured at 19w in females (only suggestive significant (p-value<1E-05) CpGs ordered by position)

Table S13B. Association of placental DNA methylation with phthalate and DINCH metabolite levels measured at 35w in females (only suggestive significant (p-value<1E-05) CpGs ordered by position)

**TableS14.**

Table S14A. Summary of results of the sensitivity model additionally adjusted for gestational age at 19w and 35w

Table S14B. Summary of results of the sensitivity model performed on European population at 19w and 35w

Table S14C. Summary of results of the sensitivity model not adjusted for cell blood composition at 19w and 35w

Table S14D. Summary of results of the sensitivity model performed on phthalate concentrations corrected for creatinine at 19w and 35w

**TableS15.**

Table S15A. Sensitivity model ajusted for gestational age: Association of placental DNA methylation with phthalate and DINCH metabolite levels measured at 19w (only suggestive significant (p-value<1E-05) CpGs ordered by position)

Table S15B. Sensitivity model ajusted for gestational age: Association of placental DNA methylation with phthalate and DINCH metabolite levels measured at 35w (only suggestive significant (p-value<1E-05) CpGs ordered by position)

**TableS16.**

Table S16A. Sensitivity model restricted to children of European origin: Association of placental DNA methylation with phthalate and DINCH metabolite levels measured at 19w (only suggestive significant (p-value<1E-05) CpGs ordered by position)

Table S15B. Sensitivity model restricted to children of European origin: Association of placental DNA methylation with phthalate and DINCH metabolite levels measured at 35w (only suggestive significant (p-value<1E-05) CpGs ordered by position)

**TableS17.**

Table S17A. Sensitivity model unadjusted for cellular composition: Association of placental DNA methylation with phthalate and DINCH metabolite levels measured at 19w (only suggestive significant (p-value<1E-05) CpGs ordered by position)

Table S17B. Sensitivity model unadjusted for cellular composition: Association of placental DNA methylation with phthalate and DINCH metabolite levels measured at 35w (only suggestive significant (p-value<1E-05) CpGs ordered by position)

**TableS18.**

Table S18A. Sensitivity model corrected for creatinine: Association of placental DNA methylation with phthalate and DINCH metabolite levels measured at 19w (only suggestive significant (p-value<1E-05) CpGs ordered by position)

Table S18B. Sensitivity model corrected for creatinine: Association of placental DNA methylation with phthalate and DINCH metabolite levels measured at 35w (only suggestive significant (p-value<1E-05) CpGs ordered by position)

**Table S19.**

Table S19A. Enriched gene-sets of phthalate and DINCH metabolites measured at 19w

Table S19B. Enriched gene-sets of phthalate and DINCH metabolites measured at 35w

**Table S20. Comparison of FDR adjusted CpGs from previous studies with results from BiSC cohort**
