## Supplementary Methods for "Association of in utero exposure to phthalate and DINCH metabolites with placental DNA methylation"

***Biological sample collection and DNA and RNA extraction***

Maternal (at 12 or 32 weeks of pregnancy or at delivery) and umbilical cord blood samples were collected in EDTA tubes (Vacutainer, Ref BD-368381). In the laboratory, EDTA tubes were centrifuged at 2000 g for 10 min and plasma, buffy coat and red cells were separated. DNA was extracted from 200 ul of buffy coat using the QIAsymphony platform and the QIAsymphony DSP DNA Mini Kit (Qiagen, Ref 937236) at the Hospital del Mar Research Institute (IMIM). DNA extraction was done separately for mothers and children, and, in both cases, samples were completely randomized.

Six-hundred and eleven placentas were collected based on maternal consent and the feasibility of collection at the hospital. Biopsies were obtained by trained gynaecologists following a harmonized protocol across hospitals. Briefly, placenta biopsies of around 2.5 cm (from the maternal to the fetal side) and 1 cm width were obtained from two opposite quadrants at a distance of around 3-4 cm from site of cord insertion. Then, these biopsies were cut in two, giving a total of 4 biopsies of 2.5 x 0.5 cm. Two of them (one from each quadrant) were directly frozen in liquid nitrogen and transferred to -80C. The other two biopsies were treated with RNAlater and then divided in four pieces of 0.5 x 0.5 cm, corresponding roughly to the fetal membranes, the upper fetal villi, the lower fetal villi and the maternal decidua. Finally, all the biopsies were stored at -80C for future use. For genomic DNA and RNA extraction, a fragment of approximately 5-6 cm^3^ (30-40 mg) was dissected from the fetal villi biopsy below the fetal membranes collected in RNAlater. Then, the tissue was disrupted/homogenized using a bead mill (bead beater). After that, 3 ul of an equimolar mix of a set of spike-ins (100 uM each) were added to the tissue homogenate to track size-selection steps of the process, as described elsewhere^13^. Genomic DNA and RNA was isolated using the AllPrep®DNA/RNA/miRNA Universal Kit, (Qiagen), and quality was evaluated on a NanoDrop spectophotomer (Thermo Scientific) and on 1% agarose gels. RNA was also evaluated for quality in a 2100 Bioanalyzer instrument (Agilent). The whole process was done in batch of 8 randomized samples, in two rounds, and finally DNA and RNA samples were stored at -80C.

***Genome-wide genotyping***

A total of 1676 DNA samples (1633 unique) were selected for genotyping at the Spanish National Genotyping Centre (CEGEN) (Spain), including 1011 maternal samples (1,002 unique) and 665 child samples (648 unique). Among the child DNA samples, 548 were obtained from umbilical cord blood and the rest from placenta. Maternal DNA samples were obtained from peripheral blood at any time point mentioned above. Genome-wide genotyping was conducted using the Infinium Global Screening (GSA) array v.3.0 with Multi-disease (MD) add-on content from Illumina, that contains 730,059 genetic variants (<https://www.illumina.com/products/by-type/microarray-kits/infinium-global-screening.html>). Before genotyping DNA samples were quantified with the ADNds Quant-iT™ PicoGreen™ kit and an aliquot of 250 ng was transferred into the genotyping plate. Samples were genotyped following the manufacturer’s recommendations in three rounds: (i) pilot study including maternal blood and cord blood DNAs, (ii) maternal blood DNAs, (iii) child blood and placenta DNAs. Two HapMap samples were included in each 96-well plate as internal controls of the quality of the whole process. Genotyping clustering was conducted using GenomeStudio GenTrain 3.0. Genetic variants were annotated in b37 + strand using the GSAMD-24v3-0-EA_20034606_A1 manifest.

Quality control of the genome-wide genetic data was conducted with PLINK v1.9 and v2 following the standard recommendations (<https://zzz.bwh.harvard.edu/plink/>, ^1^). First, 68,337 genetic variants which were not genotyped in any of the samples were filtered out. Second, we conducted the sample quality control and filtered out samples with a call rate <97% (n=15), discordant for sex (n=12), and with a heterozygosity > +- 5 standard deviations (n=4). Relatedness was estimated using the PI_HAT parameter. All mother-child pairs were identified correctly. Samples with a PI_HAT >0.25, which is indicative of 2^nd^ degree relativeness (n=54) were eliminated, including one of the intended duplicates, mothers participating twice, siblings, other relatives, as well as 16 incorrect samples. Samples without genetic consent we also excluded (n=137). Next, we conducted the genetic variant quality control and filtered out SNPs with a call rate <97% (n=14,187), with a minor allele frequency (MAF) <1% (n=151,870), not in Hardy-Weinberg equilibrium (HWE) p-value <1E-06 (n=6,290), or not annotated to any chromosome or with duplicated positions (n=1,277). Ancestry prediction was done using the GRAFpop v2.4 ^2^. Predicted ancestries were curated using the questionnaire information on self-reported ethnicity and country of birth of the parents and grandparents, ending up with the following number of participants by main ancestry groups: 1,176 European (73.9%), 391 Latin American (24.6%) and 24 Other (1.5%). We did not filter any sample based on the genetic background. Twenty GWAS principal components (PCs) were estimated for the whole population using a SNP pruning method in the PLINK tool, as well as for the subset of individuals classified as European. First two GWAS PCs explained 30.47% and 9.10% of the variance in the whole population and 2.20% and 2.00% in the Europeans, while all 20 PCs explained 72.15% and 35.4%, respectively. The final dataset consisted of 1,454 samples (947 mothers and 507 children, of which 490 were paired).

Up to 40M variants were imputed with the Haplotype Reference Consortium (HRC) version 1.1 panel ^3^ at the Sanger Institute server (<https://www.sanger.ac.uk/tool/sanger-imputation-service/>), using EAGLE2 for the phasing and PBWT for the imputation. After imputation, the following filtering of the variants was applied: MAF>1%, HWE p-value >1E-06, quality of imputation (INFO) >0.8, ending up with around 5,237,924 SNPs.

***Placental DNA methylation***

DNA methylation was assessed in 624 placental DNA samples (including 35 duplicates – 589 unique individuals) with the Infinium MethylationEPIC BeadChip from Illumina, following manufacturer’s protocol in the Human Genome facility (HUGE-F) at the Erasmus Medical Centre core facility. Briefly, 750 ng of DNA were bisulfite-converted using the EZ 96-DNA methylation kit following the manufacturer’s standard protocol, and DNA methylation measured using the Infinium protocol.

The methylation data was pre-processed using the PACEAnalysis R package (v.0.1.9) (https://www.epicenteredresearch.com/). The pre-processing pipeline consists of sample quality control, probe quality control, normalization, batch correction, estimation of cell type proportions, and winsorization of outlier values. Fifty-nine samples corresponding to 24 individuals were discarded according to: low quality (N=5), sample call rate < 95% (N=3), sex inconsistencies (N=9), samples with substantial contamination with DNA from other samples (log2 odds < -1) (N=11) (Heiss J. et al., 2018), duplicates calculated using the probes to genotype 59 single nucleotide polymorphisms (SNPs) included in the array (N=29) and siblings (N=2). Exclusion of one of the duplicate pairs was done randomly. A total of 565 samples remained after the sample quality control. Probes having a call rate < 95% were eliminated according to the SeSAME method (Zhou W., et al.2018). Dye-bias and Noob background correction, followed by normalization of the data with the functional normalization method were applied as implemented in the minfi R package (Jean-Philippe Fortin et al.2017)(Timothy J. Triche et al.2013). Then, to correct for the bias of type-2 probes values the beta-mixture quantile (BMIQ) normalization was applied (Andrew E. Teschendorff et al.2013). After that, we explored the clustering of the data through Principal Component Analysis (PCA) and tested the association of the 12 first PCs with main variables (hospital, sex, ethnic origin, birth before or during SARS-CoV-2 pandemic, maternal education and maternal smoking during pregnancy) and technical variables (plate, array, extraction batch, time to placental storage, DNA concentration, and 260/280 and 260/230 ratios). Array batch effect was controlled with the ComBat method (Johnson et al. 2007). Finally, to correct for the possible outliers, we winsorized the extreme values to the 1% percentile (0.5% in each side), where percentiles were estimated with the empirical beta-distribution. DNA methylation values are expressed as beta values, where 0 means un-methylation and 1 complete methylation.

Cell type proportions of six populations (trophoblasts, syncytiotrophoblast, nucleated red blood cell, Hofbauer cells, endothelial cells, and stromal cells) were estimated from DNA methylation using the placenta reference panel from the 3st trimester (term) implemented in the planet R package (V Yuan et al.2021).

***Identification of fetal cis mQTLs***

Fetal cis mQTLs within a window of +-0.5 Mb were identified with the TensorQTL tool ^4^. TensorQTL fits a linear regression model for each trait (DNA methylation at each CpG) and each SNP adjusting for covariates. Models were run in individuals of all ancestrie. DNA methylation was treated as beta values (from 0 to 1) / was rank-based inverse normal transformed. Covariates of the models were: child’s sex, child’s gestational age at birth, child’s ancestry based on 5 GWAS PCs, 6 placental cell type proportions estimated from the DNA methylation data, and DNA methylation PCs (mPCs). mPCs were estimated from the residuals of linear regression models being the DNA methylation the outcome and the covariates of the mQTL model (child’s sex, child’s gestational age, child’s ancestry, and 6 placental cell type proportions) the predictors. The % of variation explained by each mPC site was estimated and if >20 mPCs were needed to explain 80% of the variation, then the number of mPCs added in the models was restricted to 20. mPCs were excluded from the model if they were associated with any SNP at p-value <1E-07. None was excluded in our study. Multiple-testing correction threshold was fit to p-value <5E-08.

**Funding**

BiSC cohort has received funding from the European Research Council (ERC) under the European Union’s Horizon 2020 research and innovation programme (785994 – AirNB project), from the Health Effects Institute (4959-RFA17-1/18-1 – FRONTIER project), from the European Union’s Horizon 2020 research and innovation programme-EU.3.1.2. (874583 - ATHLETE project) and [H2020-EU.3.1.1.](https://cordis.europa.eu/programme/id/H2020-EU.3.1.1./en) (GA964827 – AURORA project), from AXA Research Fund (MOOD-COVID project), from Agence nationale de sécurité sanitaire de l'alimentation, de l'environnement et du travail (ANSES) ( 2019/01/039 - HyPAXE project), from the AGAUR-Agència de Gestió d'Ajuts Universitaris de Recerca (2017 SGR 826 - Population Neuroscience group), from the Centro de Investigación Biomédica en Red  de Epidemiología y Salud Pública   (CIBERESP) (CB06/02/0041), from the Instituto de Salud Carlos III (ISCIII) and the European Regional Development Fund (ERDF) - Maternal and Child Health and Development Network (SAMID) (RD16/0022/0014 and RD16/0022/0015), and from the Instituto de Salud Carlos III (ISCIII) and the European Union Next Generation EU - Primary Care Interventions to Prevent Maternal and Child Chronic Diseases of Perinatal and Developmental Origin Network (RICORS-SAMID) (RD21/0012/0001 and RD21/0012/0003).

Genome-wide genotyping data was funded by the Instituto de Salud Carlos III (ISCIII) and co-funded by European Union (ERDF) "A way to make Europe" (PI20/01116 – ENTENTE project) and the Centro Nacional de Genotipado-CEGEN (PRB2-ISCIII). Placental DNA methylation was funded by the Instituto de Salud Carlos III (ISCIII) and co-funded by European Union (ERDF) "A way to make Europe" (PI20/00190 – ALMA project), and from the European Joint Programming Initiative “A Healthy Diet for a Healthy Life” (JPI HDHL and Instituto de Salud Carlos III) (AC18/00006 - NutriPROGRAM project).
